# A sensitive and affordable multiplex RT-qPCR assay for SARS-CoV-2 detection

**DOI:** 10.1101/2020.07.14.20154005

**Authors:** Martin A.M. Reijns, Louise Thompson, Juan Carlos Acosta, Holly A. Black, Francisco J. Sanchez-Luque, Austin Diamond, David A. Parry, Alison Daniels, Marie O’Shea, Carolina Uggenti, Maria C. Sanchez, Alan O’Callaghan, Michelle L.L. McNab, Martyna Adamowicz, Elias T. Friman, Toby Hurd, Edward J. Jarman, Frederic Li Mow Chee, Jacqueline K. Rainger, Marion Walker, Camilla Drake, Dasa Longman, Christine Mordstein, Sophie J. Warlow, Stewart McKay, Louise Slater, Morad Ansari, Ian P.M. Tomlinson, David Moore, Nadine Wilkinson, Jill Shepherd, Kate Templeton, Ingolfur Johannessen, Christine Tait-Burkard, Jürgen G. Haas, Nick Gilbert, Ian R. Adams, Andrew P. Jackson

**Affiliations:** MRC Human Genetics Unit, MRC Institute of Genetics and Molecular Medicine, The University of Edinburgh, Edinburgh, UK; NHS Lothian, The South East of Scotland Clinical Genetic Service, Western General Hospital, Edinburgh, UK; Cancer Research UK Edinburgh Centre, MRC Institute of Genetics and Molecular Medicine, The University of Edinburgh, Edinburgh, UK; Centre for Genomic & Experimental Medicine, MRC Institute of Genetics and Molecular Medicine, The University of Edinburgh, Edinburgh, UK; Centre Pfizer-University of Granada-Andalusian Government for Genomics and Oncological Research (Genyo), Granada, Spain; Division of Infection Medicine, Edinburgh Medical School, The University of Edinburgh, Edinburgh, UK; The Roslin Institute and Royal (Dick) School of Veterinary Studies, The University of Edinburgh, Edinburgh, UK; The Milner Centre for Evolution, Department of Biology and Biochemistry, University of Bath, Bath, UK; NHS Lothian, Medical Microbiology and Virology Service, Royal Infirmary of Edinburgh, Edinburgh UK

## Abstract

With the ongoing COVID-19 pandemic, caused by the novel coronavirus SARS-CoV-2, there is need for sensitive, specific and affordable diagnostic tests to identify infected individuals, not all of whom are symptomatic. The most sensitive test involves the detection of viral RNA using RT-qPCR, with many commercial kits now available for this purpose. However, these are expensive and supply of such kits in sufficient numbers cannot always be guaranteed. We therefore developed a multiplex assay using well-established SARS-CoV-2 targets alongside a human cellular control (*RPP30*) and a viral spike-in control (PhHV-1), which monitor sample quality and nucleic acid extraction efficiency respectively. Here, we establish that this test performs as well as widely used commercial assays, but at substantially reduced cost. Furthermore, we demonstrate >1,000-fold variability in material routinely collected by nose-and-throat swabbing, and establish a statistically significant correlation between the detected level of human and SARS-CoV-2 nucleic acids. The inclusion of the human control probe in our assay therefore provides a quantitative measure of sample quality that could help reduce false negative rates. We demonstrate feasibility of establishing a robust RT-qPCR assay at ∼10% of the cost of equivalent commercial assays, which could benefit low resource environments and make high volume testing more affordable.

## Introduction

The COVID-19 pandemic, caused by the novel coronavirus SARS-CoV-2 [1], originated in Wuhan (China) in December 2019 and rapidly spread across the globe, resulting in substantial mortality [2, 3] and widespread economic damage. Until a vaccine becomes available, public health strategies centred on reducing the rate of transmission are crucial to mitigating the epidemic, for which effective and affordable testing strategies to enable widespread population surveillance are essential. The most sensitive test to diagnose infected individuals involves the detection of SARS-CoV-2 viral RNA using RT-qPCR, most commonly using samples collected with nasopharyngeal (nose-and-throat) swabs (NTS), although there is increasing evidence that the use of saliva may be a valid alternative [4-7]. Many commercial kits are now available, most of which employ multiplex RT-qPCR, detecting 2 or 3 different SARS-CoV-2 targets, and generally include an internal control to show successful nucleic acid extraction. However, such kits are often costly and their supply in sufficient numbers cannot always be guaranteed. We therefore developed a similar multiplex assay using well-established SARS-CoV-2 targets and internal controls, which can be carried out at a significantly lower cost and provides more flexibility to ensure resilience against potential shortages in reagent supplies.

Our assay makes use of the Takara One Step PrimeScript III RT-qPCR kit. This reagent was used in the first high profile publication to describe SARS-CoV-2 [1], and it has since been shown to outperform a number of similar reagents [8]. Before commercial COVID-19 assays were available, various in-house assays were published on the WHO website [9]. Based on the data available at the time (March 2020), we decided to focus our initial efforts on targeting the following SARS-CoV-2 genes (Fig 1A; [10-12]): E (envelope), RdRp (RNA-dependent RNA polymerase) and N (nucleocapsid). Corman et al (2020) proposed the E gene as a useful target for first line screening, with the RdRp gene suggested as a good target for confirmatory/discriminatory assays [13, 14]. The N gene was central to the USA Centers for Disease Control and Prevention (CDC) in vitro diagnostics emergency protocol, with three different primer/probe sets (N1, N2 and N3) used against different portions of this viral gene [9]. The CDC protocol also included a probe against human *RPP30*, a single copy gene encoding the protein subunit p30 of the Ribonuclease (RNase) P particle, to ensure the presence of a sufficient number of cells in patient samples and successful isolation of intact nucleic acids.

**Figure 1.**
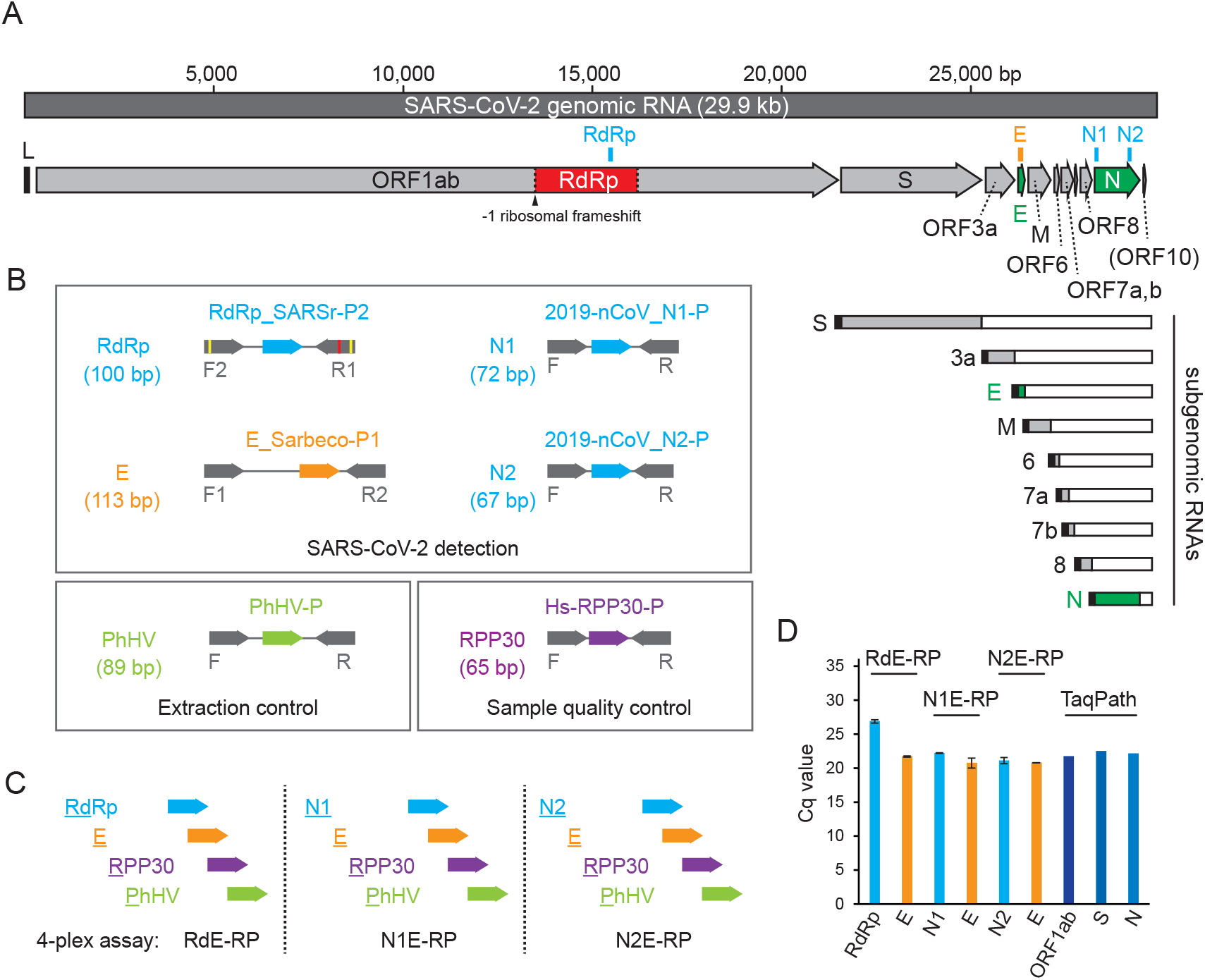
Primers and probes used in our multiplex qRT-PCR assays detect SARS-CoV-2 RNA. **A:** Location of qRT-PCR amplicons on the SARS-CoV-2 genome. Open reading frames (ORFs) and qRT-PCR target sites (orange, blue) in RdRp, E and N gene indicated. After a -1 ribosomal frameshift (arrow head) on ORF1ab of the genomic RNA, the pp1ab polypeptide is formed and RdRp/nsp12 released by proteolytic cleavage (dotted lines). Genes 3’ of ORF1ab on the positive-sense genome (including E and N) are transcribed (from negative-sense RNA) as subgenomic RNAs and include a short leader sequence (L, black box) at their 5’ ends [10]. **B:** qRT-PCR primers (grey) and probes (colour) used in this study. PCR product sizes are indicated between brackets. Yellow and red lines in RdRp primers indicate degenerate nucleotides and a mismatch respectively (as present in the original design [14]). To detect RdRp we used the RdRp_SARSr-P2 probe, which detects 2019-nCoV/SARS-CoV-2 and not SARS-CoV. The N1 and N2 assays are specific to SARS-CoV-2, whereas the E gene assay also detects SARS-CoV [9,13,14]. **C:** Probes used in the RdE-RP, N1E-RP and N2E-RP 4-plex assays detect 2 SARS-CoV-2 targets and 2 controls. **D:** Using our 4-plex assays, E, N1 and N2 give Cq values (mean ± SD, n = 2 experiments) comparable those for ORF1ab, S and N from the TaqPath assay (mean of technical triplicates, n = 1 experiment) when detecting cultured SARS-CoV-2.

In early versions of these protocols, all probes were labelled with fluorescein amidite (FAM), and separate reactions were therefore needed to detect each target. To increase efficiency, we developed a multiplex assay using 4 different fluorescent labels (FAM, HEX, CAL Fluor Red 610 and Quasar 670) for each of the probes, allowing their detection in a single reaction. In the final version of our assay, we use previously described primers and probes against the well-established SARS-CoV-2 E and N gene (N1 and N2) targets, as well as a human cellular control and a viral spike-in control (Fig 1B, C): human *RPP30* and Phocine Herpes Virus 1 (PhHV-1, hereafter referred to as PhHV). The rationale behind the human control is that a considerable number of patients with clinical and radiological signs of COVID-19 are PCR negative; and poor quality of swab samples with no or little usable patient material is one possible explanation for this [15]. In essence, the *RPP30* control provides a measure of sample quality. In addition, a defined amount of PhHV is spiked into each sample with the lysis buffer at the start of the nucleic acid isolation procedure, resulting in a known cycle quantification or Cq value (also referred to as the cycle threshold or Ct value, [16]). Detection of PhHV (using the Glycoprotein B gene as a target) simultaneously controls for extraction and amplification efficiency, and indicates absence of PCR inhibitors [17, 18].

If paired with an in-house RNA extraction protocol, our assay can be performed for less than £2 (GBP; $2.50, USD) per test, excluding cost of plastic consumables, which could mean a potential 10-fold difference in cost compared to commercial kits. Here we present data that demonstrates equivalent performance to the commercial TaqPath COVID-19 Combo Kit (CE-IVD; Thermo Fisher Scientific) and Abbott RealTime SARS-CoV-2 assay. We also document the utility of inclusion of *RPP30* as a human internal control to provide important sample quality information.

## Results

### N and E gene multiplex assays sensitively detect viral RNA

With the aim of developing a sensitive and affordable assay for the detection of SARS-CoV-2 RNA, we tested three different 4-plex strategies (Fig 1B, C). All made use of *RPP30* (HEX) and PhHV (Cy5) probes employed as internal controls (controlling for the presence of human cells in patient samples, and successful nucleic acid isolation, respectively), as well as a CFR-labelled (CAL Fluor Red 610) probe for the SARS-CoV-2 E gene. To enhance assay sensitivity and specificity, a FAM-labelled probe against a second SARS-CoV-2 target was included in each of the three assays: RdRp, N1 or N2 (N gene). We named these tests RdE-RP (<u>Rd</u>Rp, <u>E</u>, <u>R</u>PP30, <u>P</u>hHV), N1E-RP (<u>N1</u>, <u>E</u>, <u>R</u>PP30, <u>P</u>hHV) and N2E-RP (<u>N2</u>, <u>E</u>, <u>R</u>PP30, <u>P</u>hHV) respectively (Fig 1C). Initial validation tests showed that these assays were capable of detecting cultured SARS-CoV-2, with Cq values for E, N1 and N2 similar to those for ORF1ab, S and N gene obtained using the ThermoFisher TaqPath COVID-19 assay. In contrast, RdRp Cq values were substantially higher (Fig 1D; 4.4-6.1 cycles above the other targets).

Next, the RdE-RP, N1E-RP and N2E-RP assays were used to test SARS-CoV-2 positive and negative patient samples (n = 19), comparing them to the commercial TaqPath assay and Abbott RealTime SARS-CoV-2 assay (which detects RdRp and N gene). The N1E-RP and N2E-RP assays both correctly identified all 9 samples that had tested positive using the TaqPath and Abbott assays (Table 1). The RdE-RP assay performed less well, identifying 7 of these samples correctly, giving inconclusive results for the other two (P18 and 19), with E gene but not RdRp detected. Overall, we find RdRp detection to be at least 20-fold less sensitive than for E gene, N1 and N2 under our assay conditions; consistent with reports by others [19]. This may be due to a mismatch in the reverse primer employed in the RdRp (P2) assay, as originally designed [14]. Both N1E-RP and N2E-RP assays also identified positive samples that scored negative with the commercial tests, suggesting potentially higher sensitivity of our assays. Of the 10 patient samples that were negative for the Abbott assay, 9 were similarly shown to be negative using the N1E-RP assay, whereas 8 of these were negative for the N2E-RP assay. Patient 11, previously negative using the Abbott assay, was inconclusive with TaqPath (1 of 3 SARS-CoV-2 targets detected) and N2E-RP assays (1 of 2 targets detected), but positive in the N1E-RP assay. Patient 12 had previously tested negative using both Abbott and TaqPath assays, and was also negative for N1E-RP; however, this sample tested weakly positive for both SARS-CoV-2 targets in the N2E-RP assay. Cq values were high for both P11 and P12, close to the limit of detection, but with multiple viral targets detected, these likely represent true positives. However, differentiating between samples with low viral loads and false positives is challenging. Analysis of such samples by Sanger sequencing of PCR products, or nanopore sequencing of RNA present could provide useful information. Further clinical evaluation and repeat sampling of the patient involved may also be a beneficial route to a secure clinical diagnosis.

**Table 1.**
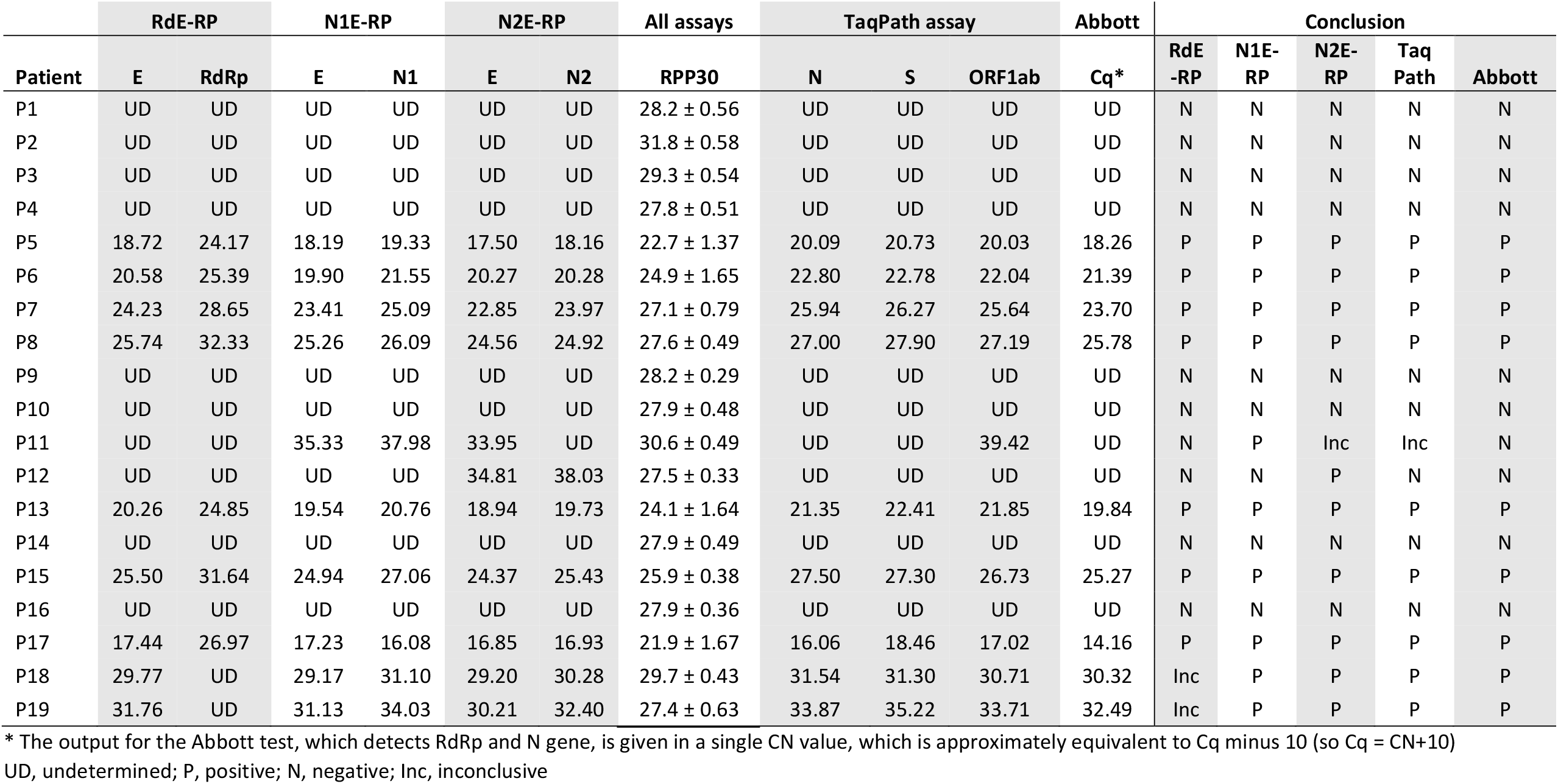
The multiplex assay detecting RdRp and E gene (RdE-RP) is not sufficiently sensitive; assays detecting N and E gene (N1E-RP, N2E-RP) are.

As our initial characterisation demonstrated the N1E-RP and N2E-RP assays to be at least as sensitive as two commercial assays, TaqPath COVID-19 Combo Kit (Thermo Fisher Scientific) and Abbott RealTime SARS-CoV-2 (Abbott Laboratories), we focussed on these two assays for further validation experiments.

### Multiplex assays for N and E genes can detect between 1 and 50 copies of IVT RNA

To determine the detection limit for the N1E-RP and N2E-RP assays, in vitro transcribed (IVT) RNA controls for each of the SARS-CoV-2 targets were prepared. An equimolar mix was used to make a dilution series (for 10,000 down to 1 copy of RNA per reaction; a 50-copy control was also included) and Cq values determined in triplicate using both assays (Fig 2A). To test our nucleic acid isolation protocol, all dilutions after extraction were simultaneously tested in triplicate (Fig S1A). All probes (E, N1 and N2) reproducibly detected 50 copies (Fig 2A, Fig S1A and Table S1). The N1 probe detected 10 copies reproducibly (6 out of 6; 6/6), while the N2 probe did so in some reactions (4/6) of the respective 4-plex assays. The E probe detected 10 copies reproducibly in the N2E-RP assay (6/6), but only did so in half of the N1E-RP reactions (3/6). As might be expected, single copies of RNA were only detected in a small proportion of reactions for each probe: E (2/12), N1 (3/6) and N2 (1/6). We therefore conclude that our assays have the sensitivity to detect between 1 and 50 copies of IVT RNA (Fig 2A, Fig S1A and Table S1), both pre and post-extraction.

**Figure 2.**
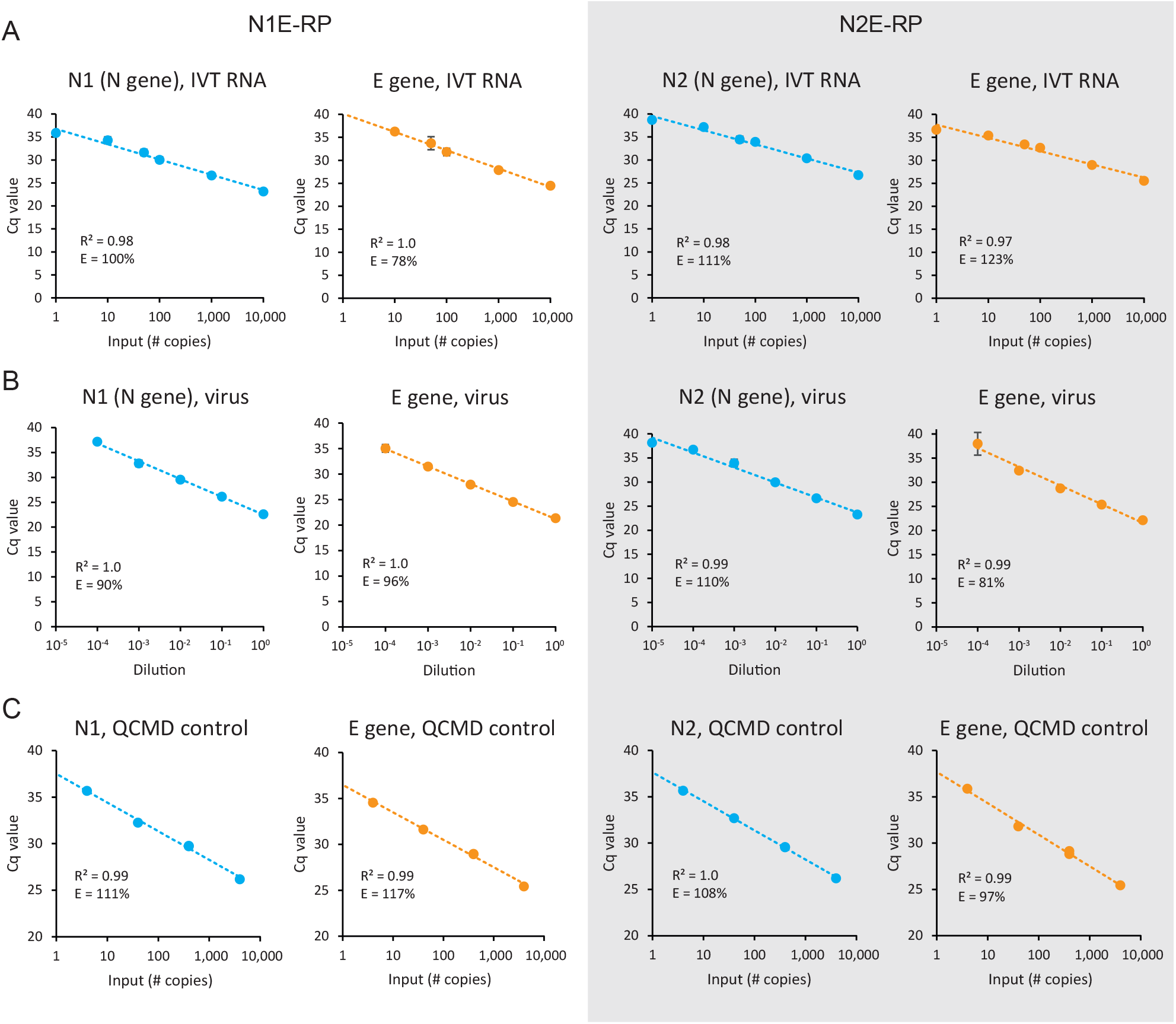
The N1E-RP and N2E-RP 4-plex assays detect between 1 and 10 copies of SARS-CoV-2 RNA. N1E-RP and N2E-RP RT-qPCR assays were performed with the following controls **A:** 1 to 10,000 copies of SARS-CoV-2 control RNA (IVT, in vitro transcribed); **B:** Serial dilu on of RNA isolated from cultured SARS-CoV-2 (hCoV-19/England/02/2020). Mean ± SD for technical triplicates shown for A and B. Also, see Table S1 and S2. **C:** RNA isolated from QCMD viral controls (BetaCoV/Munich/ChVir984/2020). These controls contained different amounts of SARS-CoV-2 virus at known copy number (see Materials and Methods for details; also see Table S3). R2 values for logarithmic trend line fitting; E, amplifca on efficiency (see Materials and Methods).

### N1E-RP and N2E-RP assays can detect between 1 and 3 copies of viral RNA

To confirm the range of detection for total viral RNA, nucleic acids isolated from cultured SARS-CoV-2 were used to make a dilution series (10^−1^ to 10^−6^), and Cq values determined in triplicate using the N1E-RP and N2E-RP assays (Fig 2B). To test the impact of the nucleic acid isolation procedure on extraction of low copy numbers of viral RNA, and to test the PhHV spike in control, the dilution series was also re-extracted and used for RT-qPCR simultaneously (Fig S1B). Sensitivity of detection for these samples was highest for E gene, followed by N1 and N2 (Fig 2B, Fig S1B and Table S2). Signal was lost for the 10^−5^ dilution in most cases, consistent with the Cq values of the undiluted sample (21.3-23.4) and the 100,000-fold reduction in copy number for this dilution (theoretically predicted Cq values, ∼38-40). For all extractions and RT-qPCR replicates the signal for the PhHV spike in was highly reproducible (Table S2), with a Cq value of 32.5 ± 0.40 (mean ± SD, range 30.7-33.0), indicating robust extraction efficiency and absence of PCR inhibitors.

Finally, 8 quality control samples obtained from QCMD (an international external quality assessment organisation) were also tested using the N1E-RP, N2E-RP and TaqPath assays. Each assay gave the same outcome, consistent with data provided by QCMD (Table S3), identifying 5 samples as positive (4 to 4,000 copies of SARS-CoV-2 per reaction) and 3 as negative (containing transport medium or different corona viruses). All probes used in the N1E-RP and N2E-RP assays displayed a linear range of detection down to 4 copies of viral RNA (Fig 2C). These calibration curves were retrospectively used to calculate the detection limit of our own viral RNA serial dilution (Fig 2B), showing that in these experiments our assays detected between 1 and 3 copies of genomic SARS-CoV-2 RNA.

### N1E-RP and N2E-RP multiplex assays correctly identify positive patient samples

Next, to further establish assay reproducibility in a diagnostics context, the N1E-RP and N2E-RP assays were performed on an additional 89 patient samples, and results compared to the TaqPath assay. The patient samples contained both SARS-CoV-2 positives and negatives, and were tested blind. Internal controls were included to provide confirmation of successful nucleic acid extraction and absence of PCR inhibitors, with lysis buffer spiked with both MS2 (an RNA bacteriophage that infects *Escherichia coli*) and PhHV (a DNA virus that infects seals), detected by the TaqPath and N1E-RP/N2E-RP assays respectively. In addition, the same three controls were performed for each assay: an extracted viral transport medium control (negative for SARS-CoV-2 and *RPP30*, positive for PhHV), a non-extracted water only control (negative for all targets) and a non-extracted in vitro transcribed RNA positive control (50 copies; positive for SARS-CoV-2, negative for *RPP30* and PhHV).

Results for controls were as anticipated (Table S4), with signal absent (undetermined) for SARS-CoV-2 and *RPP30* targets for the negative controls, and Cq values for the SARS-CoV-2 RNA positive control (50 copies) similar to those obtained previously (Fig 2A). The PhHV control gave consistent Cq values for both N1E-RP (32.5 ± 1.1) and N2E-RP assays (33.3 ± 1.2; Table S4, Fig S2A), confirming reliable and reproducible extraction of nucleic acids from patient samples; similar to the MS2 control used in the TaqPath assay (mean Cq value, 25.6 ± 0.9; Table S4, Fig S2A). Out of the 89 samples, the TaqPath assay identified 75 samples as negative, 1 as inconclusive and 13 as positive. Both the N1E-RP and N2E-RP assay detected the same 13 positive samples, and the majority of TaqPath negative samples were similarly negative in our assays (n = 74). For the N1E-RP assay 6 of the negative samples had Cq values between 39.0 and 43.2 for N1 (E gene not detected), suggesting potentially higher sensitivity of the N1 probe in this assay. The sample that was inconclusive with the TaqPath assay (P75) was positive for both N1E-RP and N2E-RP assays, consistent with this being a true positive. In addition, there was one sample (P53) that was negative with TaqPath, but positive for both N1E-RP and N2E-RP assays, albeit with very high Cq values (between 35.7 and 39.2), close to the limit of detection.

Altogether, our data (for n = 108 patient samples) establish that the sensitivity of the N1E-RP and N2E-RP assays is similar to, if not higher than, the TaqPath assay (Fig 3).

**Figure 3.**
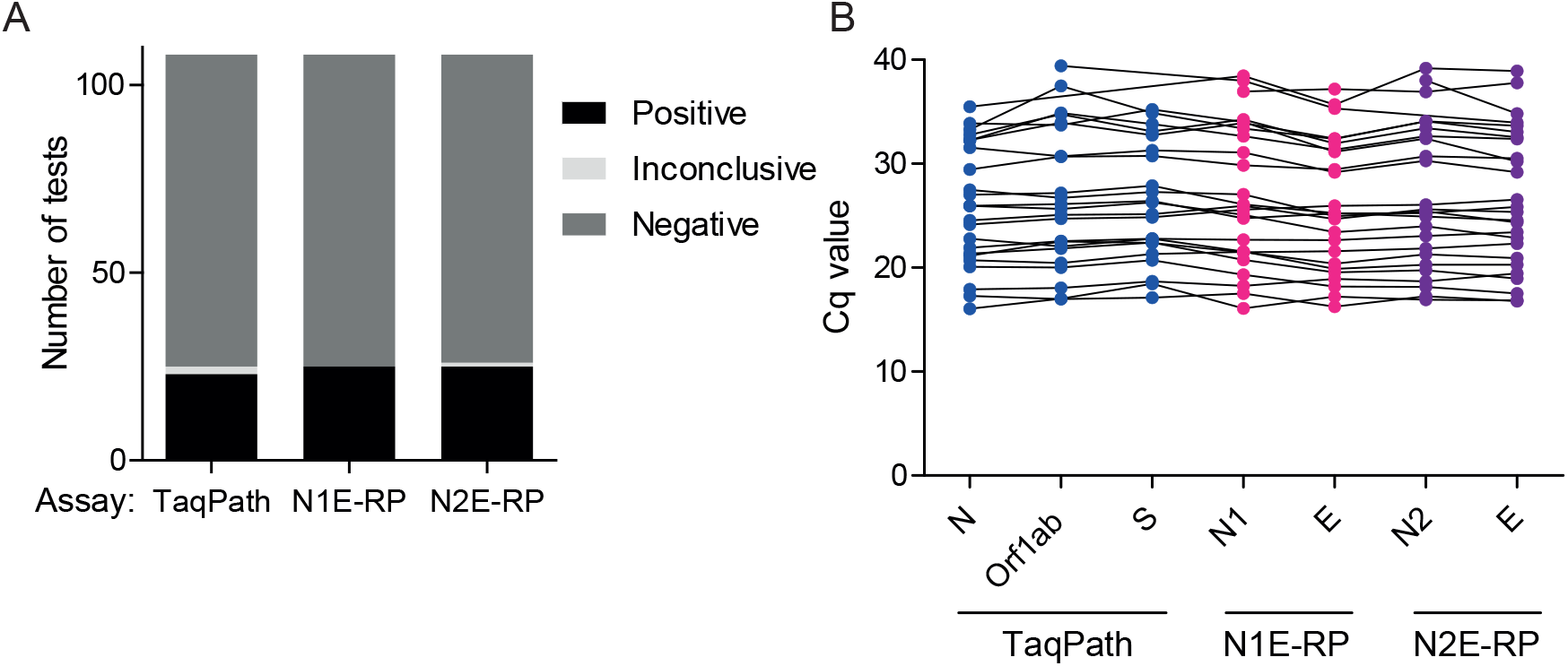
The N1E-RP and N2E-RP 4-plex assays perform similarly to the TaqPath assay, correctly identifying positive and negative patient samples. **A:** The Taqpath, N1E-RP and N2E-RP assays each identified a similar numbers of positives and negatives among 108 patient samples. Inconclusive: only one of the SARS-CoV-2 targets was detected. **B:** Cq values for each of the SARS-CoV-2 targets in the TaqPath (N, Orf1ab, S), N1E-RP (N1, E) and N2E-RP (N2, E) were comparable (for n=24-26 positive patients). Also, see Table 1 and S4.

### Substantial variability in NTS quality, as measured by human *RPP30*, impacts on assay sensitivity

The range of Cq values for the human *RPP30* control was much greater than that of the PhHV internal control (Table S4, Fig 4A, B and Fig S2). This indicated that there was considerable variability in the amount of cellular material present in different patient samples. The *RPP30* primer/probe set has good amplification efficiency and was able to detect 10 copies of positive control nucleic acids (Fig S3), hence Cq values for this probe represent a good measure of the presence of intact cellular nucleic acids in patient samples. Although *RPP30* was detected in all samples, Cq values ranged from 20.1 to 31.7 for the N1E-RP assay and from 20.3 to 32.1 for the N2E-RP assay, which equates to a difference of between 3,100 and 3,600-fold in extracted nucleic acids between the best and worst samples. Although the distribution of positives (Fig 4A, B) shows that samples with high *RPP30* Cq values can still test positive, this is not an adequate measure of the likelihood of false negatives among samples with similarly low levels of human material. A statistically significant linear correlation between Cq values for each of the viral probes (E, N1, and N2) and the Cq values for the *RPP30* sample quality probe (p < 0.001; Fig 4C and data not shown) established that samples containing fewer human cells are more likely to have less SARS-CoV-2, decreasing the chance of detection. To visually demonstrate the impact of this, we normalised SARS-CoV-2 Cq values to the sample with the lowest amount of human material detected (reference sample, *RPP30* Cq value 31.7/32.1). Due to the linear correlation between *RPP30* and viral Cq values, subtracting the difference in *RPP30* Cq between a particular positive sample and the reference sample from the SARS-CoV-2 Cq gives an indication of what the viral Cq may have been had it contained the same low amount of human material present in the reference sample. This shows that of the 26 positive samples we detected, between 4 and 6 (15-23%) would not have tested positive, with at least one of the viral targets exceeding the detection limit (Cq > 40; Fig 4D and Fig S4A). Theoretically, using this approach, even a strong positive sample (SARS-CoV-2 Cq value of 28.2) of good quality (*RPP30* Cq value of 20.3) may have given a false negative test result (SARS-CoV-2 Cq value of 40) if it had contained the same low amount of human material as the reference sample (*RPP30* Cq value of 32.1; viral Cq: 32.1-20.3+28.2=40). Conversely, normalising samples to an optimal quality sample (RPP30 Cq 20.1/20.3) gives an indication of what viral Cq values may have been if all samples had contained a similar (more optimal) amount of material (Fig 4E, Fig S4B). This highlights the possibility that a proportion of apparent SARS-CoV-2 negative samples are in fact false negatives as a result of insufficient material in the swab fluid. Notably, the SARS-CoV-2 Cq values clustered more strongly after normalisation (Fig 4D, E; Fig S4). This reduced variability not only shows that the amount of human material present in NTS samples impacts on assay sensitivity, but also suggests that variability in viral load is not as great as implied by RT-qPCR data without normalisation.

**Figure 4.**
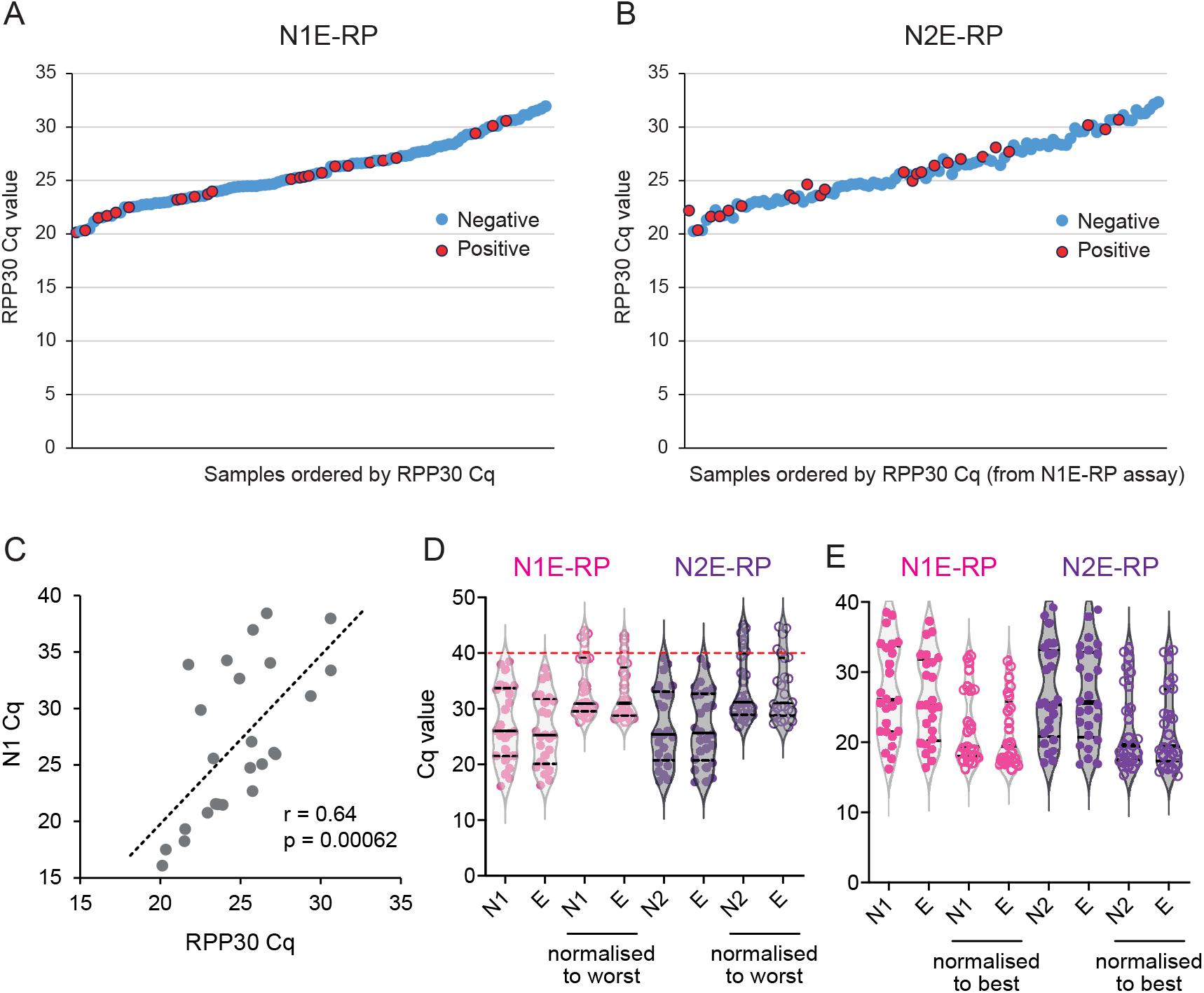
The *RPP30* control indicates substantial variability in sample quantity/quality, impacting on assay sensitivity. **A, B:** *RPP30* Cq values for 108 patient samples ranked from low to high (based on N1E-RP ranks) for N1E-RP and N2E-RP assays. SARS-CoV-2 positives, marked in red, are detected for samples with low as well as high RPP30 Cq values. **C:** *RPP30* and SARS-CoV-2 Cq values are highly correlated, demonstrating that samples with fewer human cells have lower levels of SARS-CoV-2, supporting the validity of *RPP30* Cq as a measure of sample quality. r, Pearson’s correlation coefficient; p-value calculated by F-test. **D, E:** Normalisation to *RPP30* levels increases clustering of viral Cq values. Reduced variability in apparent SARS-CoV-2 levels when normalising to the worst (highest *RPP30* Cq; **D**) as well as the best (lowest *RPP30* Cq; **E**) sample. Plots in panel **D** demonstrate the impact of sample quality on assay sensitivity, with 4-6 positives (15-23%) below the detection limit (above the red line, viral Cq > 40) for a worst quality sample scenario.

## Discussion

Here, we describe a user-friendly protocol for an accurate and affordable SARS-CoV-2 RT-qPCR test. Although we did not detect substantial differences between our two assays, others have reported higher sensitivity of the N1 over the N2 assay [19]. We therefore recommend the use of the N1E-RP assay for primary testing, and the N2E-RP assay could be employed if initial results are inconclusive. We provide detailed materials and methods to enable others to rapidly set up this assay in their own laboratory or to adapt it to locally available equipment and reagents. While we provide extensive validation of the reagents and instruments used to perform these multiplex RT-qPCR assays, our methods allow some flexibility. Probes with different labels as well as alternative Real-Time PCR machines could be used, as long as the different dyes can be detected simultaneously. Also, spike-in controls other than PhHV could be used, such as recombinant DNA or RNA. However, virus would better mimic extraction of SARS-CoV-2 RNA, and other viruses with an RNA genome (e.g. lentivirus, routinely produced in many molecular biology laboratories) would make particularly good controls, not only confirming successful RNA extraction, but also controlling for RNA stability and reverse transcription. As a further development, replacing E gene with M or S gene probes, could provide N2M or N2S assays as fully independent second line tests. Both M and S gene assays have been shown to have high sensitivity [20, 21] and their target regions display low sequence variation (Table S5). Additional improvements to our protocol could include the use of control primers/probe specific to a human RNA transcript (the *RPP30* primers/probe described here detect both RNA and genomic DNA) as this would ensure that samples contain intact RNA [22]. However, it should be stressed that any changes to the protocol may also change the sensitivity of SARS-CoV-2 detection, and new protocols should undergo appropriate validation before use for diagnostic purposes.

Our assays have high analytical sensitivity, equivalent to commercial CE-IVD kits. RT-qPCR tests are molecular tests with high intrinsic accuracy, however false positive and false negative results can occur. The use of multiplex assays that detect multiple SARS-CoV-2 targets, such as those reported here, reduces the chance of both. Off-target reactivity is one possible cause of false positives, and although some have reported high false positive rates for the E gene assay [20, 23], this does not match our experience. In two patients, our N1E-RP and N2E-RP assays detected virus, albeit weakly, whereas commercial assays did not. As multiple SARS-CoV-2 targets were positive, these are likely true positive results and not due to off-target reactivity. False positives can also occur due to lab issues such as sample mislabelling, data entry errors, reagent contamination with target nucleic acids or contamination of primary specimens. However high standards of quality control at all stages of testing, and effective mitigation strategies should quickly identify problems. Additionally, sample re-testing with an independent assay and/or patient re-sampling should also be effective measures to counter false positives, particularly in low pre-test probability situations such as mass screening. False negative test results are an important ongoing issue, estimated to be somewhere between 2 and 54% [24-26]. Sequence variation at primer/probe binding sites could be one factor resulting in false negatives. However, for the primers and probes used the chance of this is low (>97.6% of 97,782 strains have no relevant changes; Table 2, Table S5), and strains with mutations in two independent targets should be very rare. Therefore sequence variation is not expected to be a significant contributing factor to the number of false negatives. In contrast, low sample quality provides a much more likely explanation; and this may be particularly important in case of self-sampling. Systematic inclusion of a human control to provide sample quality metrics, could therefore have utility in reducing the number of false negatives. Testing saliva, as an alternative to NTS sampling, could also be beneficial as a modality that may have less-sample to sample variability [7].

**Table 2.** Percentage of known SARS-CoV-2 genomic sequences with mutations in primer/probe binding sites for E gene, N1 and N2 assays.

| Assay | Primer/probe | Percentage of strains with any mismatch/deletion* |  | Percentage of strains with mismatch in 5 most 3' nt* |  |
| --- | --- | --- | --- | --- | --- |
|  |  | Per primer/probe | Per assay** | Per primer | Per assay*** |
| E gene | E_Sarbeco-F1 | 0.10% | 0.26% | 0.006% | 0.007% |
|  | E_Sarbeco-P1 | 0.12% |  | (0.008%) |  |
|  | E_Sarbeco-R2 | 0.04% |  | 0.001% |  |
| N1 | 2019-nCoV_N1-F | 0.29% | 2.39% | 0.034% | 0.157% |
|  | 2019-nCoV_N1-P | 1.79% |  | (0.057%) |  |
|  | 2019-nCoV_N1-R | 0.33% |  | 0.124% |  |
| N2 | 2019-nCoV_N2-F | 0.35% | 1.82% | 0.153% | 0.169% |
|  | 2019-nCoV_N2-P | 0.33% |  | (0.008%) |  |
|  | 2019-nCoV_N2-R | 1.19% |  | 0.015% |  |

High quality genome sequences analysed only (n=97,782).
\* 0-0.003% of genomes have deletions in primer/probe binding regions, so majority are mismatches. Presence of these changes does not necessarily impact on primer/probe performance. Mutations at the 3' ends for the primer regions (here defined as the final 5 nt) are more likely to affect assay sensitivity.
\*\* Mismatches/deletions in the primer/probe set used in single target assay
\*\*\* Mismatches in the forward and reverse primers only. For probes values are given between brackets for reference (in the "per primer" column).

Absence of *RPP30* signal (undetected or Cq >40) clearly indicates that absence of viral detection cannot be interpreted as a negative test result and that a repeat test is required (Table S6). However, utilising *RPP30* Cq values when interpreting an apparent SARS-CoV-2 negative sample requires further consideration: what should the *RPP30* Cq limit be for which to order a repeat test? One option would be to simply set an arbitrary cut-off, e.g. one could decide to re-test any samples with *RPP30* Cq >30, or with Cq values above the 95^th^ centile (Cq ∼ 31 for our 108 samples). To determine robust cut-off limits, collection of *RPP30* data for a much larger number of patient samples would be desirable. This would allow development of diagnostic algorithms that could incorporate a sample quality score based on the level of *RPP30* detected. Nonetheless, *RPP30* data, even as it stands, are useful for the interpretation of cases for which only one of the SARS-CoV-2 targets is (weakly) positive, with samples with high *RPP30* Cq values interpreted with particular caution. In such cases, repeat testing of the same sample (with an independent assay of equal or better sensitivity) would be advisable, and repeat patient specimen collection and testing might also be considered (see Table S6 for guidance). Ultimately, the clinical sensitivity of any diagnostic test is influenced by multiple factors, including sample timing relative to symptom onset, sample type and sample quality. The inclusion of a human control in our assays provides an internal sample quality control, supporting improved interpretation of test results, which could contribute to reducing false negative rates.

Taken together we show that sensitive and robust RT-qPCR assays for the detection of SARS-CoV-2 are available at a fraction of the cost of comparable commercial assays. The use of these assays could make widespread population testing more feasible.

## Materials and Methods

### Patient samples

Samples were collected from symptomatic individuals by trained healthcare professionals using combined nose-and-throat swabbing, and processed for diagnostic testing using validated CE-IVD assays. Excess samples were then used to validate the in-house multiplex assays, with specimen anonymization by coding, compliant with Tissue Governance for the South East Scotland Scottish Academic Health Sciences Collaboration Human Annotated BioResource (reference no. SR1452). A variety of swabs and viral transport media (VTM) were used. In each case, swabs were placed in VTM and kept at ambient temperature until processed (within 24 h).

### Nucleic Acid isolation

Nucleic acids were isolated using the Omega Mag-Bind Viral DNA/RNA 96 Kit (Cat. No. M6246), following the Supplementary Protocol for NP Swabs (April 2020 version). Briefly, 200 μl VTM was taken from patient swab sample inside a Class-2 safety cabinet and mixed with 240 μl TNA Lysis Buffer, 1 μl carrier RNA and extraction controls (MS2, provided as part of the TaqPath COVID-19 Combo Kit, and PhHV, provided by the laboratory of Jürgen Haas) in screw capped tubes, for virus inactivation. After incubation at room temperature for at least 15 min, samples were transferred from tubes into 96-well KingFisher Deep well plates (Cat. No. 95040450) containing 280 μl isopropanol and 2 μl Mag-Bind Particles per well, using a Biomek NX^P^ Automated Liquid Handler (Beckman Coulter). Plates were then moved and the isolation completed on a KingFisher Flex robot (Cat. No. 5400610) as instructed by the manufacturer, including washes with 350 μL VHB Buffer and 2x 350 μL SPR Buffer, and RNA finally eluted in 50 μl of nuclease-free water in KingFisher 96 microplates (Cat. No. 97002540). Also see Supplementary Material, Protocol 2.

An in-house version of a magnetic bead-based isolation was tested and shown to perform similarly in preliminary experiments (data not shown). The use of this protocol could further reduce the cost per test, but would require additional validation. For details of this RNA isolation protocol, see Supplementary Material, Protocol 2.

### Primers and probes

Primers and probes (Table 3) were synthesised and HPLC purified by LGC BioSearch Technologies (Risskov, Denmark), and dissolved in IDTE (10 mM Tris, 0.1 mM EDTA, pH 8.0) to prepare 100 μM stocks. Pre-prepared primer/probe mixes (FAM-labelled) for N1, N2 and *RPP30* were obtained from Integrated DNA Technologies (IDT, USA; Cat. No. 10006713). Since we developed our assay, N1, N2 and *RPP30* primers and probes also became available from IDT as 100 μM stocks, but can also be purchased from other reputable oligonucleotide synthesis companies. All nucleic acid stocks and dilutions were prepared in Eppendorf DNA LoBind tubes (Cat. No. 10051232).

**Table 3.**
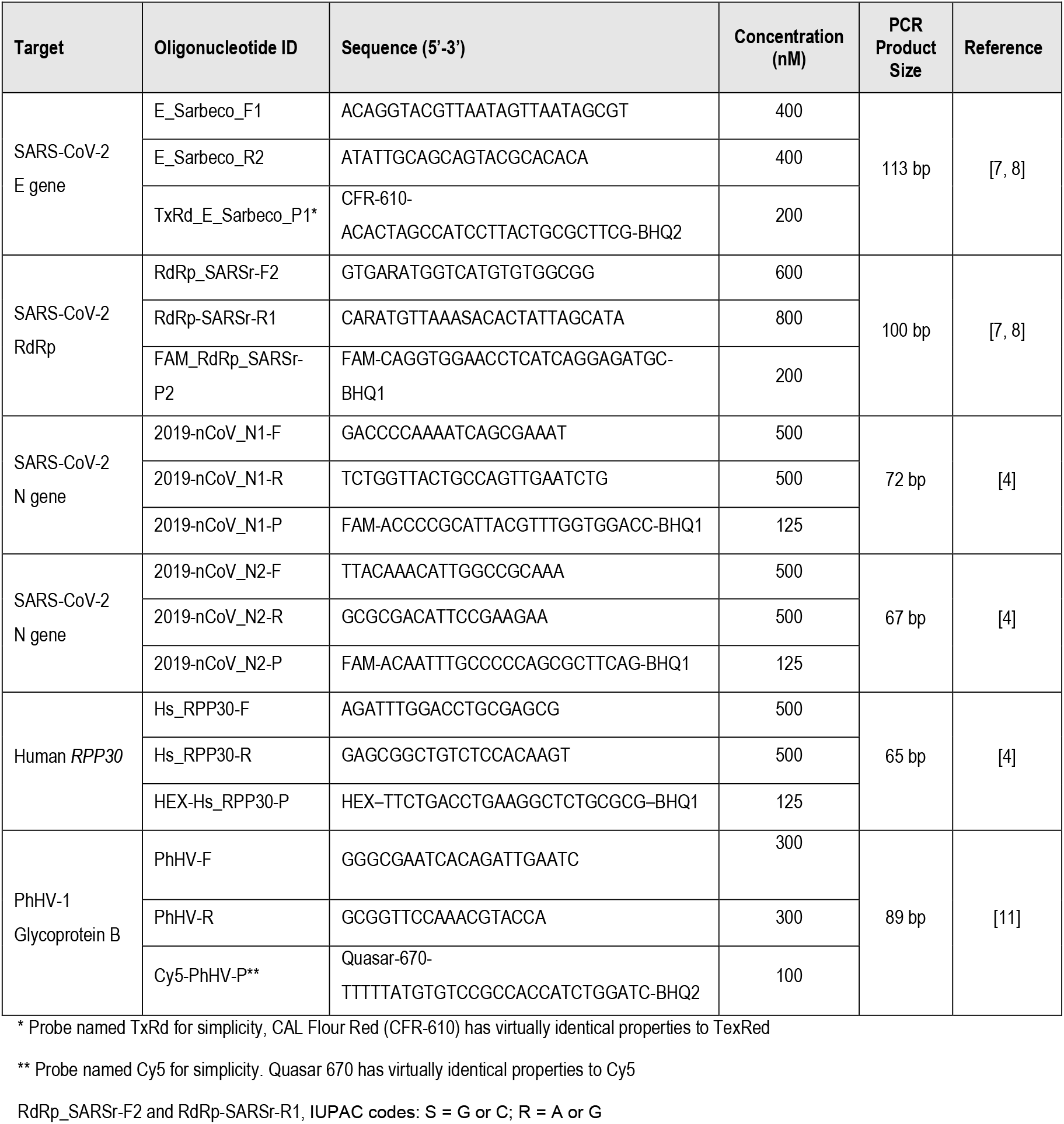
Primer/Probe details for 4-plex assays.

Primer/probe mixes (50x) were prepared for E gene (20 μM E_Sarbeco_F1, 20 μM E_Sarbeco_R2, 10 μM TxRd_E_Sarbeco_P1), RdRp (30 μM RdRp_SARSr-F2, 40 μM RdRp_SARSr-R2, 10 μM FAM_RdRp_SARSr-P2), *RPP30* (25 μM Hs_RPP30-F, 25 μM Hs_RPP30-R, 6.25 μM HEX-Hs_RPP30-P) and PhHV (15 μM PhHV-F, 15 μM PhHV-R, 5 μM Cy5-PhHV-P). The N1 and N2 primers/probes were purchased premixed (∼13.3x) from IDT. These individual primer/probe mixes, were then used to prepare a single mix for each of the 4-plex assays: 12.5x for RdE-RP (with equal volumes of each of the relevant mixes), 7.4x for N1E-RP and N2E-RP (with equal volumes of the E, *RPP30* and PhHV mixes, combined with 3.7x volumes of N1 or N2 mix). Mixes were stored at -20°C, with working stocks kept at 4°C.

Primers and probes included in the TaqPath COVID-19 Combo Kit (Thermo Fisher Scientific, Cat. No. A47814) detect SARS-CoV-2 ORF1ab, N and S gene; those in the Abbott RealTime SARS-CoV-2 assay (Cat. No. 09N77-090) detect RdRp and N gene. Further details are not available, as this information is proprietary.

### RT-qPCR

All RT-qPCRs were performed on Applied Biosystems 7500 Fast Real-Time PCR Systems with ABI 7500 software v2.3, using MicroAmp Fast Optical 0.1 mL 96-well reaction plates (Cat. No. 4346906) and Optical Adhesive film (Cat. No. 4311971). For our assays we used the Takara One Step PrimeScript III RT-qPCR kit (Cat. No. RR600B). These were compared to the TaqPath COVID-19 Combo Kit (Thermo Fisher Scientific, Cat. No. A47814) and the Abbott RealTime SARS-CoV-2 assay (Cat. No. 09N77-090), used as instructed by the manufacturer. The TaqPath assay was performed on the ABI 7500 Fast System and the Abbott assay was performed on the M2000 system. Experiments using the 4-plex assay were performed as described below, with a user-friendly protocol provided in Supplementary Material, Protocol 1.

Reaction master mixes were prepared (20 μl per reaction) for each assay, before adding 5 μl of template RNA per reaction, brief centrifugation and starting the PCR program. For the RdE-RP 4-plex assay, per reaction 12.5 μl of One-Step mix, 5.5 μl of nuclease-free water, 2 μl of 12.5x primer/probe mix and 5 μl of template RNA were mixed. For the N1E-RP and N2E-RP 4-plex assays, per reaction 12.5 μl of One-Step mix, 4.16 μl of nuclease-free water, 3.34 μl of 7.4x primer/probe mix and 5 μl of template RNA were mixed. For all 4-plex reactions the PCR program was: 5 min at 52°C, 10 s at 95°C, then 45 cycles of 3 s at 95°C and 30 s at 60°C. For detection the FAM, JOE, TEXAS RED and CY5 channels were used.

### Amplification efficiency

Amplification efficiency (E) was determined for all standard curves using the slope of the linear curve fit when plotting Cq values versus the log of input amounts. An E of 100% means that the number of molecules of the target sequence double during each replication cycle. The following formula was used: E = 100 x (10^−1/slope^ – 1).

### Positive controls

Positive control RNAs generated by in vitro transcription were provided by Sylvie Behillil (Institut Pasteur, Paris, France) for E gene [9] and by Christine Tait-Burkard (Roslin Institute, Edinburgh, UK) for RdRp [13, 14], N1/N3 and N2 [9]. An equimolar mix of all RNAs was prepared at 2.5 × 10^8^ copies/μl, and aliquots stored at -80°C. Dilution series were prepared in nuclease-free water, in Eppendorf DNA LoBind tubes (Cat. No. 10051232), at 2,000, 200, 20, 10, 2 and 0.2 copies/μl.

A cultured SARS-CoV-2 control (strain hCoV-19/England/02/2020; GISAID Accession EPI_ISL_407073, [27-29]) was provided by Rory Gunson (NHS Molecular Development in Virology and Microbiology, Glasgow, UK). Except for the reported S15519T mismatch in RdRp-SARSr_R1 [19], this strain has no mutations in target sites for the primers and probes used in our assays [30].

### QCMD controls

Quality Control for Molecular Diagnostics (Glasgow, UK) provided controls as part of the “QCMD 2020 Coronavirus Outbreak Preparedness (CVOP) EQA Pilot Scheme” [31]. RNA extractions were performed using 200 μl of each sample, eluting in 50 μl. After samples were tested blind with our assays, expected results along with sample identities were provided by QCMD. Quantification of control samples was carried out by QCMD prior to distribution within the EQA scheme, using droplet digital PCR (ddPCR) with E-gene primers and probe [13, 14] on the Biorad droplet digital PCR platform. A serial dilution of inactivated SARS-CoV-2 (strain BetaCoV/Munich/ChVir984/2020; GenBank Accession MT270112, [32]) was prepared and each dilution replicate tested 4 times using both RT-qPCR and ddPCR assays. Regression analysis was used to assess the linearity across the dilution series, and the analytical measurement range established for both assays, comparing results of each by Bland-Altman difference plot. Except for the reported S15519T mismatch in RdRp-SARSr_R1 [19], this strain has no mutations in target sites for the primers and probes used in our assays [30]. Concentrations determined by ddPCR in Log10 dPCR copies/ml were used to calculate the number of copies of input RNA.

### Statistical analysis

To determine whether a linear relationship exists between the observed Cq values for viral probes and the *RPP30* (sample quality) probe, we used Pearson’s correlation coefficient and linear regression. Linear regression p-values were calculated using an F-test. Qualitative visual model diagnostics indicated in each case that the statistical assumptions of linear regression models were not violated; in particular the normality and homoscedasticity of residuals. Statistical analysis was performed using R version 4.0.3 [33].

### Primer/probe mismatch analysis

SARS-CoV-2 (hCoV-19) genome sequences and multiple sequence alignment (MSA) of 131,759 strains were downloaded from the GISAID EpiCov™ database [27, 29]. Local alignments were generated between each oligo and the hCoV-19/Wuhan/WIV04/2019 reference strain using biopython’s pairwise2 module [34]. Alignment coordinates were then transposed to the corresponding positions in the MSA. For each strain sequence in the MSA, mismatches and gaps were counted; where mixed bases (IUPAC ambiguity codes with the exception of N) were present in either oligo or strain sequence a position was considered to “match” if there was overlap between the mixed bases. If either oligo sequence or strain sequence contained gaps relative to one another a pairwise local alignment was performed between the oligo and the strain sequence corresponding to the oligo position ± 20 flanking nucleotides in order to detect any ungapped matches between strain sequence and oligo.

To ensure only high-quality sequences were included in the analysis, genome sequences with >1% Ns or with gaps ≥ 95^th^ percentile were excluded. Sequences with Ns in oligo regions were also excluded, leaving a total of 97,782 sequences. Code used to count mismatches between strains and oligos is provided at https://github.com/david-a-parry/SARS-CoV-2_oligos_vs_strains.

## Supporting information

Supplemental file - primers & probes on SARS-CoV-2 genome (SnapGene file)

## Data Availability

n/a

## Acknowledgments

We thank Sylvie Behillil (Institut Pasteur, Paris, France), Rory Gunson (NHS Molecular Development in Virology and Microbiology, Glasgow, UK), and Paul Wallace (QCMD, Glasgow, UK) for reagents and information; and Angela Ingram, Derek Mills, Maggie Arbuckle, Joyce Begbie, Heather Coupar, Eilidh Guild, Samantha Griffiths, Garry Jempson, Alain Kemp, Frances Rae, and Thomas Williams for support. CT-B and MO’S were funded by BBSRC Institute Strategic Programme grant funding (BBS/E/D/20002172, BBS/E/D/20002173). This work was supported by a UK Medical Research Council Human Genetics Unit core grant (MRC, U127580972) and by NHS National Services Scotland, as part of a technology development exercise at the NHS-led University of Edinburgh/NHS Lothian COVID-19 testing centre at the Institute of Genetics and Molecular Medicine.

## Author contributions

MAMR, APJ, conceived the project. MAMR, LT, JCA, ADi, ADa, KT, JGH, NG, designed experiments. MAMR, LT, JCA, HAB, NG, performed RNA extractions and RT-qPCR experiments. ADa, MCS, performed viral inactivation experiments. FJS-L, CU, MA, ETF, TH, EJJ, FLMC, JKR, MW, CD, DL, CM, SJW, SM, LS, MA, DM, provided technical assistance. MAMR, LT, JCA, HAB, DAP, ADa, AO’C, analysed data. MO’S, MLLM, NW, JS, KT, CT-B, JGH, provided reagents. MAMR, DM, KT, CT-B, JGH, IRA, APJ, supervised the work. IPMT, DM, NW, IJ, IRA, APJ, provided administrative organisation. MAMR, APJ, wrote the manuscript. All authors had the opportunity to edit the manuscript and approved the final manuscript.

## Supplementary information

Supplementary material includes 4 figures, 6 tables, 2 protocols and 1 additional file.

## Supplementary material

**Figure S1.**
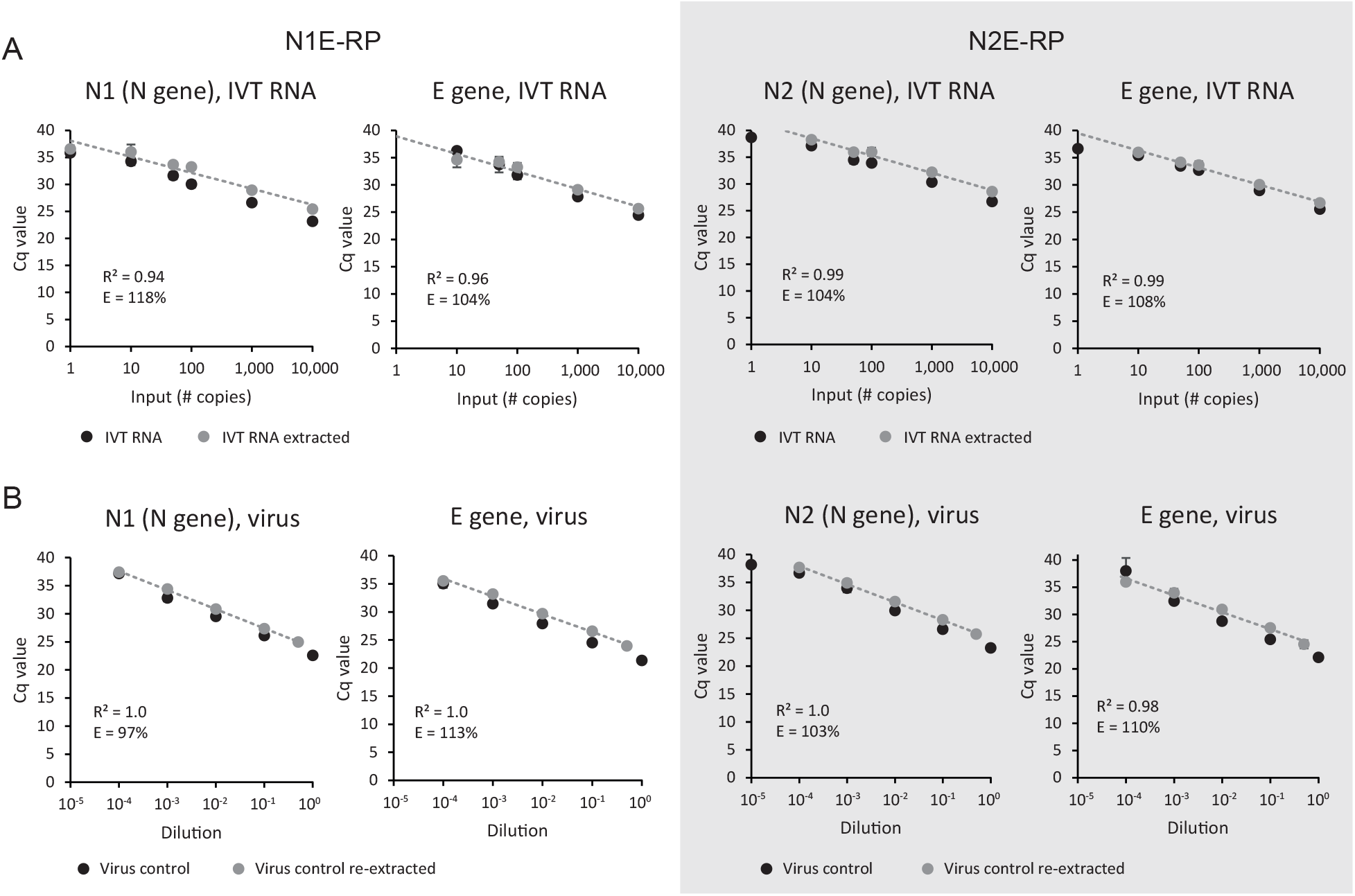
RNA extraction has no substantial impact on the sensitivity of the N1E-RP and N2E-RP 4-plex assays. N1E-RP and N2E-RP RT-qPCR assays were performed on **A:** 1 to 10,000 copies of SARS-CoV-2 control RNA (IVT, in vitro transcribed) before (as Fig 2A) and after nucleic acid extraction; **B:** a serial dilution of RNA isolated from cultured SARS-CoV-2, before (as Fig 2B) and after re-extraction. Mean ± SD for technical triplicates; R2 values for logarithmic trend line fitting and amplification efficiencies (E) for samples after (re)extraction. Also, see Table S1 and S2.

**Figure S2.**
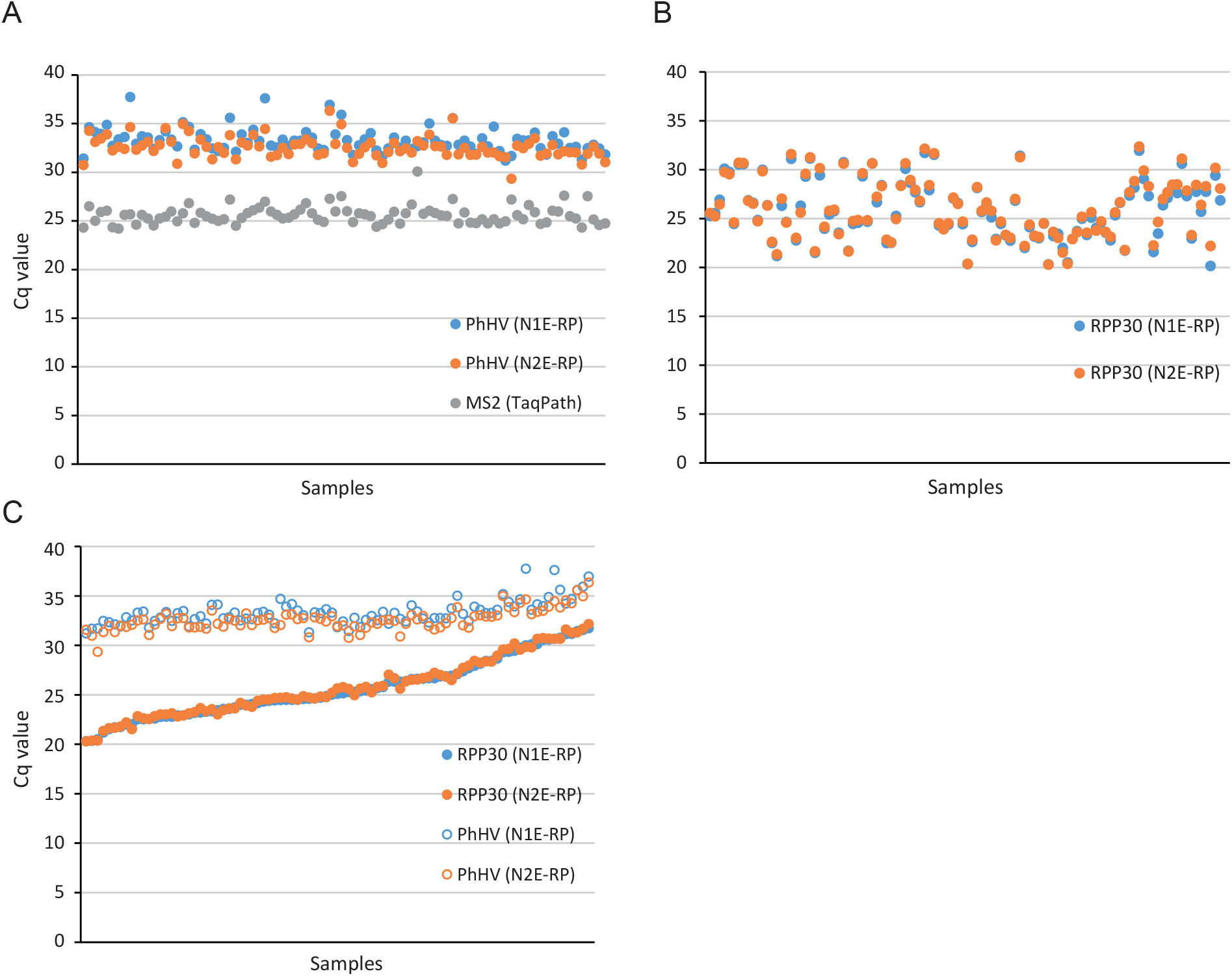
High reproducibility for extraction controls, but high variability for the human *RPP30* control in NTS samples. **A, B:** Cq values for internal controls, MS2 for TaqPath and PhHV for N1E-RP and N2E-RP assays (A), and *RPP30* controls (B). **C:** Cq values for PhHV and *RPP30* controls for N1E-RP and N2E-RP assays, ranked by *RPP30* values from the N1E-RP assay, confirm that variability does not substantially correlate with extraction efficiency.

**Figure S3.**
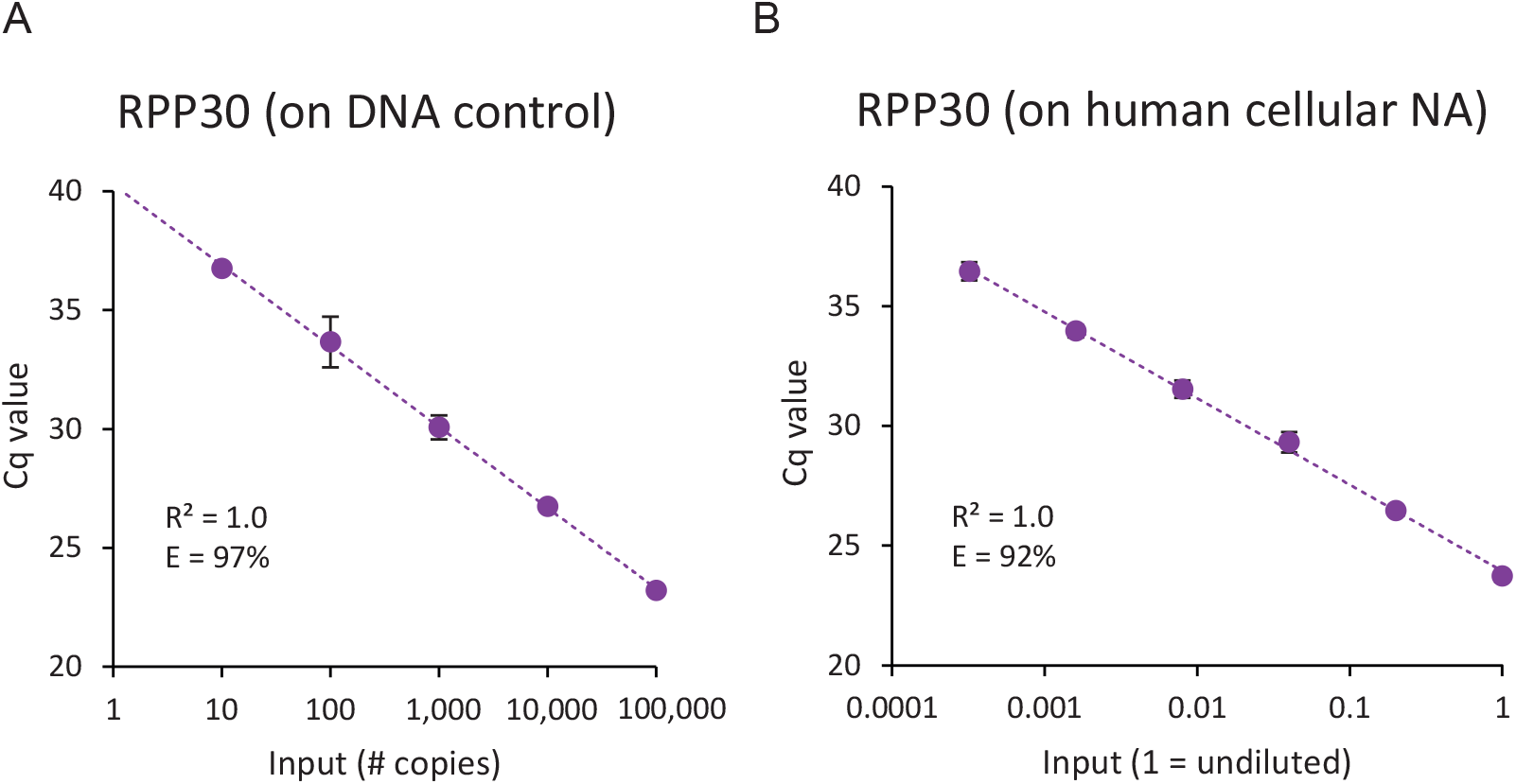
The human *RPP30* control probe can detect 10 copies of control DNA. **A:** Cq values for *RPP30* on a serial dilution of positive control plasmid DNA (100,000 down to 10 copies were tested). **B:** Cq values for *RPP30* on nucleic acids (NA) isolated from human cultured cells (1 = undiluted) and NA isolated from a serial dilution of the same cell suspension show a strong linear correlation and 92% primer efficiency. Negative control samples did not show any amplification. Data points and error bars, mean ± SD (n = 2 technical replicates). R^2^ values for logarithmic trend line fitting; E, amplification efficiency.

**Figure S4.**
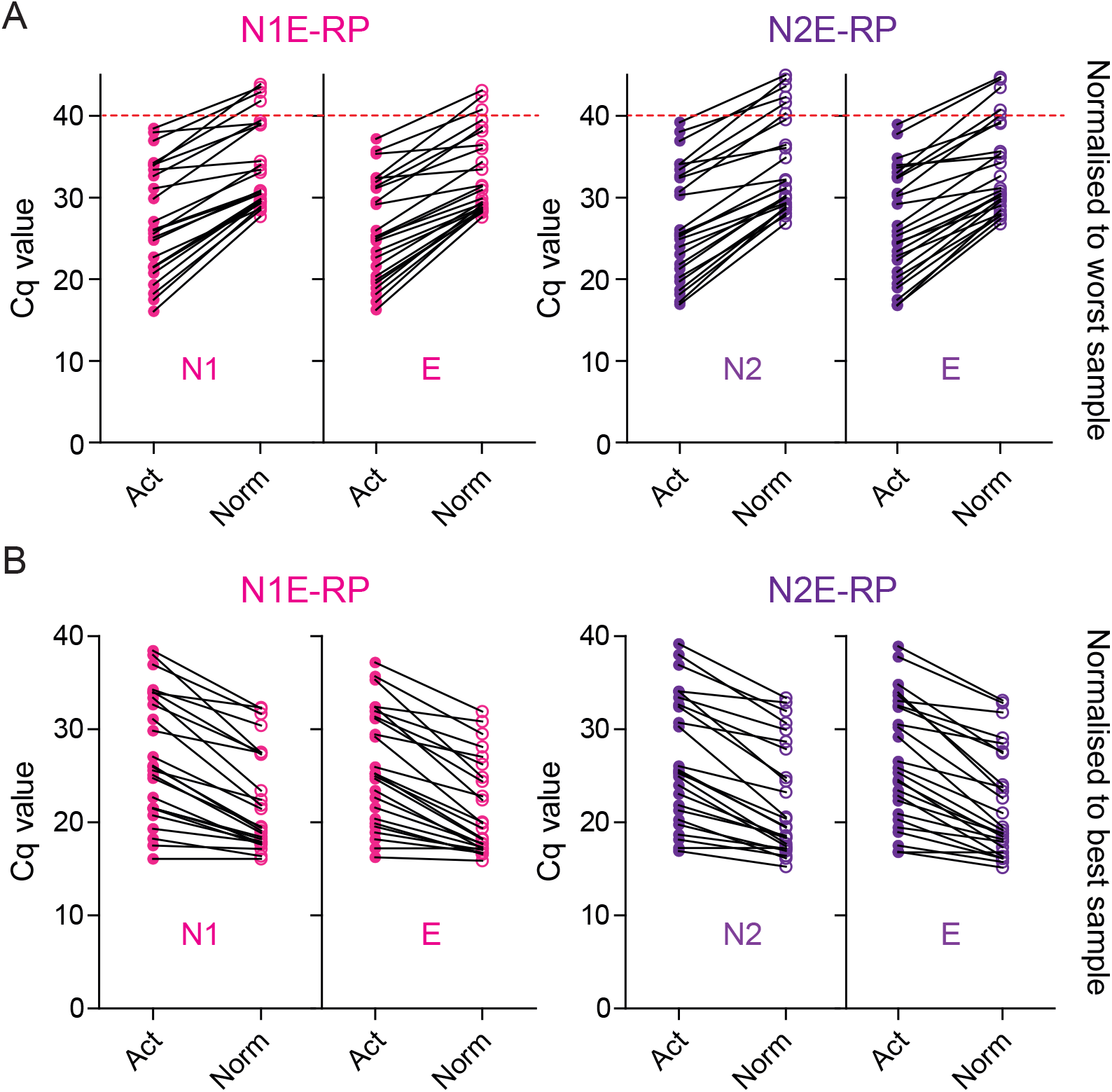
Increased chance of false negatives for low quality NTS samples, with high *RPP30* Cq values. **A, B:** Comparison of actual (Act) and *RPP30*-normalised (Norm) Cq values for SARS-CoV-2 targets shows that values cluster more strongly after normalisation, both when normalising to the worst (highest *RPP30* Cq; **A**) and best (lowest *RPP30* Cq; **B**). Increased clustering (i.e. reduced variability) demonstrates the impact of the correlation between quantity of human and viral nucleic acids in NTS samples (Fig 4C). This shows 1) that sample quality substantially impacts on assay sensitivity and 2) that variability in viral loads is smaller than non-normalised data suggest. Plots in **A** demonstrate that 4-6 positives (15-23%) would likely have been below the detection limit (above the red line, viral Cq > 40) in a worst quality sample scenario.

**Table S1.** N1E-RP and N2E-RP assay Cq values for SARS-CoV-2 RNA controls (1 to 10,000 copies) pre- and post-extraction (values used for Fig 2A and Fig S1A)

| # copies | 10,000 | 1,000 | 100 | 50 | 10 | 1 |
| --- | --- | --- | --- | --- | --- | --- |
| **N1, N1E-RP assay pre-extraction** |  |  |  |  |  |  |
| Rep 1 | 22.83 | 26.49 | 30.30 | 31.42 | 33.49 | 35.87 |
| Rep 2 | 23.36 | 26.62 | 30.50 | 31.05 | 34.43 | Undetermined |
| Rep 3 | 23.30 | 26.77 | 29.32 | 32.38 | 34.94 | Undetermined |
| Mean | 23.17 | 26.62 | 30.04 | 31.62 | 34.29 | 35.87 |
| **E gene, N1E-RP pre-extraction** |  |  |  |  |  |  |
| Rep 1 | 24.26 | 27.98 | 31.37 | 33.60 | 36.26 | Undetermined |
| Rep 2 | 24.74 | 27.73 | 32.73 | 35.22 | Undetermined | Undetermined |
| Rep 3 | 24.43 | 27.90 | 31.34 | 32.36 | Undetermined | Undetermined |
| Mean | 24.48 | 27.87 | 31.81 | 33.73 | 36.26 | Undetermined |
| **N2, N2E-RP assay pre-extraction** |  |  |  |  |  |  |
| Rep 1 | 26.63 | 30.49 | 33.99 | 34.63 | 37.34 | 38.72 |
| Rep 2 | 26.88 | 29.99 | 34.01 | 35.06 | Undetermined | Undetermined |
| Rep 3 | 26.70 | 30.58 | 33.79 | 37.34 | 36.99 | Undetermined |
| Mean | 26.74 | 30.35 | 33.93 | 34.47 | 37.16 | 38.72 |
| **E gene, N2E-RP assay pre-extraction** |  |  |  |  |  |  |
| Rep 1 | 25.50 | 28.71 | 32.84 | 33.37 | 35.25 | Undetermined |
| Rep 2 | 25.70 | 28.71 | 33.05 | 33.24 | 35.93 | 36.64 |
| Rep 3 | 25.50 | 29.56 | 32.29 | 33.85 | 35.05 | Undetermined |
| Mean | 25.57 | 28.99 | 32.73 | 33.49 | 35.41 | 36.64 |
| **N1, N1E-RP assay post-extraction** |  |  |  |  |  |  |
| Rep 1 | 25.64 | 29.02 | 33.14 | 33.59 | 35.09 | Undetermined |
| Rep 2 | 25.14 | 28.80 | 33.41 | 33.58 | 35.37 | 36.14 |
| Rep 3 | 25.50 | 29.01 | 33.22 | 33.87 | 37.62 | 37.04 |
| Mean | 25.43 | 28.94 | 33.25 | 33.68 | 36.03 | 36.59 |
| **E gene, N1E-RP assay post-extraction** |  |  |  |  |  |  |
| Rep 1 | 25.99 | 29.24 | 33.26 | 35.01 | 35.59 | Undetermined |
| Rep 2 | 25.39 | 28.88 | 34.04 | 33.87 | 33.64 | Undetermined |
| Rep 3 | 25.61 | 29.22 | 32.47 | 33.81 | Undetermined | Undetermined |
| Mean | 25.66 | 29.11 | 33.26 | 34.23 | 34.61 | Undetermined |
| **N2, N2E-RP assay post-extraction** |  |  |  |  |  |  |
| Rep 1 | 28.69 | 32.39 | 36.86 | 36.01 | 38.35 | Undetermined |
| Rep 2 | 28.33 | 31.69 | 35.75 | 35.57 | 38.17 | Undetermined |
| Rep 3 | 28.79 | 32.56 | 35.40 | 36.21 | Undetermined | Undetermined |
| Mean | 28.60 | 32.21 | 36.01 | 35.93 | 38.26 | Undetermined |
| **E gene, N2E-RP assay post-extraction** |  |  |  |  |  |  |
| Rep 1 | 26.72 | 29.89 | 33.85 | 33.40 | Undetermined | Undetermined |
| Rep 2 | 26.45 | 30.01 | 33.38 | 34.53 | 35.87 | Undetermined |
| Rep 3 | 26.96 | 30.30 | 33.79 | 34.56 | 36.10 | 36.40 |
| Mean | 26.71 | 30.07 | 33.67 | 34.16 | 35.98 | 36.40 |

**Table S2.** N1E-RP and N2E-RP assay Cq values for cultured SARS-CoV-2 dilution series (before and after re-extraction; values used for Fig 2B and Fig S1B)

|  | neat | 10<sup>-1</sup> | 10<sup>-2</sup> | 10<sup>-3</sup> | 10<sup>-4</sup> | 10<sup>-5</sup> | 10<sup>-6</sup> |
| --- | --- | --- | --- | --- | --- | --- | --- |
| **N1, N1E-RP assay** |  |  |  |  |  |  |  |
| 1 | 22.53 | 26.46 | 29.42 | 32.21 | 37.19 | UD | UD |
| 2 | 22.83 | 26.03 | 29.87 | 32.79 | UD | UD | UD |
| 3 | 22.40 | 25.96 | 29.35 | 33.48 | 37.16 | UD | UD |
| Mean | 22.59 | 26.15 | 29.55 | 32.83 | 37.18 | UD | UD |
| **E gene, N1E-RP assay** |  |  |  |  |  |  |  |
| 1 | 21.31 | 24.77 | 27.97 | 31.35 | 34.22 | UD | UD |
| 2 | 21.38 | 24.38 | 28.03 | 31.50 | 35.27 | UD | UD |
| 3 | 21.35 | 24.41 | 27.84 | 31.55 | 35.63 | UD | UD |
| Mean | 21.35 | 24.52 | 27.94 | 31.46 | 35.04 | UD | UD |
| **N2, N2E-RP assay** |  |  |  |  |  |  |  |
| 1 | 23.08 | 26.76 | 29.95 | 33.56 | 37.21 | 38.06 | UD |
| 2 | 23.36 | 26.64 | 29.79 | 34.81 | 36.50 | 38.29 | UD |
| 3 | 23.32 | 26.46 | 30.14 | 33.49 | 36.44 | UD | UD |
| Mean | 23.25 | 26.62 | 29.96 | 33.95 | 36.72 | 38.17 | UD |
| **E gene, N2E-RP assay** |  |  |  |  |  |  |  |
| 1 | 22.00 | 25.48 | 28.78 | 32.41 | 36.34 | UD | UD |
| 2 | 22.22 | 25.42 | 28.57 | 32.59 | UD | 40.63 | UD |
| 3 | 22.15 | 25.31 | 28.87 | 32.31 | 39.68 | UD | UD |
| Mean | 22.12 | 25.40 | 28.74 | 32.44 | 38.01 | 40.63 | UD |
|  | 0.5 | 10<sup>-1</sup> | 10<sup>-2</sup> | 10<sup>-3</sup> | 10<sup>-4</sup> | 10<sup>-5</sup> | 10<sup>-6</sup> |
| **N1, N1E-RP assay after re-extraction** |  |  |  |  |  |  |  |
| 1 | 24.95 | 27.48 | 30.79 | 34.27 | UD | UD | UD |
| 2 | 24.93 | 27.54 | 31.08 | 34.77 | 37.44 | UD | UD |
| 3 | 25.04 | 27.12 | 30.69 | 34.26 | UD | UD | UD |
| Mean | 24.97 | 27.38 | 30.85 | 34.44 | 37.44 | UD | UD |
| **E gene, N1E-RP assay after re-extraction** |  |  |  |  |  |  |  |
| 1 | 23.85 | 26.56 | 29.63 | 33.49 | UD | UD | UD |
| 2 | 23.86 | 26.53 | 29.85 | 33.30 | UD | UD | UD |
| 3 | 24.18 | 26.59 | 29.59 | 32.77 | 35.54 | UD | UD |
| Mean | 23.96 | 26.56 | 29.69 | 33.19 | 35.54 | UD | UD |
| **N2, N2E-RP assay after re-extraction** |  |  |  |  |  |  |  |
| 1 | 25.90 | 28.27 | 31.84 | 35.46 | 37.68 | UD | UD |
| 2 | 25.76 | 28.36 | 31.78 | 34.97 | UD | UD | UD |
| 3 | 25.53 | 28.27 | 31.13 | 34.36 | UD | UD | UD |
| Mean | 25.73 | 28.30 | 31.58 | 34.93 | 37.68 | UD | UD |
| **E gene, N2E-RP assay after re-extraction** |  |  |  |  |  |  |  |
| 1 | 24.86 | 27.37 | 30.73 | 33.22 | UD | UD | UD |
| 2 | 25.00 | 27.55 | 30.81 | 34.54 | UD | UD | UD |
| 3 | 23.65 | 27.62 | 31.25 | 34.19 | 35.96 | UD | UD |
| Mean | 24.51 | 27.51 | 30.93 | 33.99 | 35.96 | UD | UD |

**Table S3.** N1E-RP, N2E-RP and TaqPath assays all correctly identify COVID-19/SARS-CoV-2 positive QCMD quality control samples.

| QCMD Sample | Content* | Log10 dPCR Copies/ml** | N1E-RP assay |  |  | N2E-RP assay |  |  | TaqPath assay |  |  | Conclusion |  |  |
| --- | --- | --- | --- | --- | --- | --- | --- | --- | --- | --- | --- | --- | --- | --- |
|  |  |  | N1 | E | PhHV | N2 | E | PhHV | N | ORF1ab | S | N1E-RP | N2E-RP | Taq Path |
| CVOP20S2-01 | SARS-CoV-2 | 4.30 | 29.72 | 28.98 | 32.22 | 29.58 | 29.14 | 32.98 | 28.06 | 28.21 | 28.15 | P | P | P |
| CVOP20S2-02 | Coronavirus – NL63 | 4.64 | UD | UD | 33.54 | UD | UD | 32.79 | UD | UD | UD | N | N | N |
| CVOP20S2-03 | SARS-CoV-2 | 3.30 | 32.26 | 31.62 | 32.66 | 32.70 | 31.8 | 32.52 | 30.24 | 29.22 | 30.02 | P | P | P |
| CVOP20S2-04 | Coronavirus – OC43 | 4.03 | UD | UD | 32.64 | UD | UD | 33.01 | UD | UD | UD | N | N | N |
| CVOP20S2-05 | Transport medium | - | UD | UD | 32.54 | UD | UD | 32.45 | UD | UD | UD | N | N | N |
| CVOP20S2-06 | SARS-CoV-2 | 4.30 | 29.77 | 28.94 | 32.90 | 29.56 | 28.82 | 32.67 | 27.96 | 27.79 | 27.63 | P | P | P |
| CVOP20S2-07 | SARS-CoV-2 | 5.30 | 26.18 | 25.42 | 31.64 | 26.20 | 25.44 | 31.93 | 24.63 | 24.52 | 24.65 | P | P | P |
| CVOP20S2-08 | SARS-CoV-2 | 2.30 | 35.69 | 34.55 | 33.02 | 35.66 | 35.88 | 32.74 | 35.89 | 36.62 | 36.14 | P | P | P |

UD, undetermined; P, positive; N, negative
\* {Matheeussen, 2020 #17} "SARS-CoV-2 strain BetaCoV/Munich/ChVir984/2020 provided by the Charité-Universitätsmedizin Berlin Institute of Virology, Berlin, Germany; human coronaviruses HCoV-NL63 and HCoV-OC43 cultivated by the University Medical Center Groningen, Groningen, the Netherlands, and sample specifications selected based on past performance data from regular provided coronavirus EQA schemes; negative control sample contained transport medium only (which was used as sample matrix for the EQA samples)."
\*\* Values obtained using ddPCR assay (see Materials and Methods for details). {Matheeussen, 2020 #17}: "Samples CVOP20S-07, -01, -03 and -08 are in a calibrated dilution series. CVOP20S-06 is a duplicate sample of CVOP20S-01. The values provided are for reference only."

**Table S4.** The N1E-RP and N2E-RP COVID-19 4-plex assays perform as well as the TaqPath CE-IVD assay.

|  | N1E-RP assay |  |  |  | N2E-RP assay |  |  |  | TaqPath assay |  |  |  | Conclusion |  |  |
| --- | --- | --- | --- | --- | --- | --- | --- | --- | --- | --- | --- | --- | --- | --- | --- |
| Patient | N1 | E | RPP30 | PhHV | N2 | E | RPP30 | PhHV | N | ORF1ab | S | MS2 | N1E-RP | N2E-RP | TaqPath |
| P20 | UD | UD | 25.56 | 30.74 | UD | UD | 25.25 | 31.41 | UD | UD | UD | 24.31 | N | N | N |
| P21 | UD | UD | 29.53 | 34.27 | UD | UD | 29.76 | 34.64 | UD | UD | UD | 26.50 | N | N | N |
| P22 | UD | UD | 24.59 | 33.10 | UD | UD | 24.47 | 34.14 | UD | UD | UD | 25.01 | N | N | N |
| P23 | UD | UD | 30.71 | 33.47 | UD | UD | 30.53 | 33.93 | UD | UD | UD | 25.90 | N | N | N |
| P24 | UD | UD | 30.67 | 33.89 | UD | UD | 30.57 | 34.87 | UD | UD | UD | 26.04 | N | N | N |
| P25 | UD | UD | 26.81 | 32.22 | UD | UD | 26.86 | 32.71 | UD | UD | UD | 24.34 | N | N | N |
| P26 | UD | UD | 26.52 | 32.60 | UD | UD | 26.60 | 33.39 | UD | UD | UD | 24.22 | N | N | N |
| P27 | UD | UD | 24.72 | 32.38 | UD | UD | 24.83 | 33.63 | UD | UD | UD | 25.62 | N | N | N |
| P28 | UD | UD | 29.83 | 34.63 | UD | UD | 29.98 | 37.73 | UD | UD | UD | 25.68 | N | N | N |
| P29 | UD | UD | 25.23 | 32.30 | UD | UD | 25.55 | 32.94 | UD | UD | UD | 24.59 | N | N | N |
| P30 | 39.97 | UD | 26.45 | 32.73 | UD | UD | 26.92 | 33.71 | UD | UD | UD | 25.62 | N | N | N |
| P31 | UD | UD | 29.77 | 33.12 | UD | UD | 30.10 | 33.58 | UD | UD | UD | 25.23 | N | N | N |
| P32 | UD | UD | 26.34 | 32.17 | UD | UD | 26.36 | 32.39 | UD | UD | UD | 24.51 | N | N | N |
| P33 | UD | UD | 24.60 | 32.81 | UD | UD | 24.63 | 33.31 | UD | UD | UD | 25.25 | N | N | N |
| P34 | UD | UD | 31.59 | 34.52 | UD | UD | 31.11 | 34.22 | UD | UD | UD | 25.43 | N | N | N |
| P35 | UD | UD | 23.00 | 33.15 | UD | UD | 22.77 | 33.35 | UD | UD | UD | 25.97 | N | N | N |
| P36 | UD | UD | 25.61 | 30.87 | UD | UD | 26.31 | 32.66 | UD | UD | UD | 24.95 | N | N | N |
| P37 | UD | UD | 29.57 | 34.96 | UD | UD | 29.28 | 35.14 | UD | UD | UD | 25.78 | N | N | N |
| P38 | UD | UD | 31.26 | 34.22 | UD | UD | 31.14 | 34.67 | UD | UD | UD | 26.81 | N | N | N |
| P39 | UD | UD | 21.62 | 31.94 | UD | UD | 21.50 | 32.33 | UD | UD | UD | 24.79 | N | N | N |
| P40 | UD | UD | 30.16 | 33.34 | UD | UD | 29.42 | 33.91 | UD | UD | UD | 25.82 | N | N | N |
| P41 | UD | UD | 22.61 | 32.60 | UD | UD | 22.49 | 33.39 | UD | UD | UD | 25.45 | N | N | N |
| P42 | UD | UD | 21.32 | 31.32 | UD | UD | 21.16 | 32.45 | UD | UD | UD | 25.23 | N | N | N |
| P43 | UD | UD | 27.02 | 32.57 | UD | UD | 26.31 | 32.17 | UD | UD | UD | 25.01 | N | N | N |
| Patient | N1 | E | RPP30 | PhHV | N2 | E | RPP30 | PhHV | N | ORF1ab | S | MS2 | N1E-RP | N2E-RP | TaqPath |
| P44 | 34.26 | 31.95 | 24.16 | 31.99 | 33.42 | 32.56 | 23.96 | 32.50 | 32.77 | 34.76 | 33.12 | 25.17 | P | P | P |
| P45 | 39.35 | UD | 30.63 | 33.82 | UD | UD | 30.75 | 35.61 | UD | UD | UD | 27.19 | N | N | N |
| P46 | UD | UD | 21.65 | 31.33 | UD | UD | 21.68 | 32.11 | UD | UD | UD | 24.49 | N | N | N |
| P47 | UD | UD | 24.73 | 33.07 | UD | UD | 24.46 | 33.90 | UD | UD | UD | 25.31 | N | N | N |
| P48 | UD | UD | 24.83 | 32.74 | UD | UD | 24.55 | 33.01 | UD | UD | UD | 25.82 | N | N | N |
| P49 | UD | UD | 29.62 | 33.85 | UD | UD | 29.32 | 34.36 | UD | UD | UD | 26.10 | N | N | N |
| P50 | UD | UD | 24.81 | 32.67 | UD | UD | 24.68 | 33.23 | UD | UD | UD | 26.37 | N | N | N |
| P51 | UD | UD | 30.65 | 34.44 | UD | UD | 30.64 | 37.60 | UD | UD | UD | 26.99 | N | N | N |
| P52 | 25.95 | 24.70 | 27.20 | 31.59 | 25.63 | 25.35 | 26.66 | 32.75 | 24.55 | 25.08 | 25.14 | 25.97 | P | P | P |
| P53 | 36.96 | 37.19 | 25.79 | 31.76 | 36.93 | 37.77 | 25.43 | 32.55 | UD | UD | UD | 25.63 | P | P | N |
| P54 | UD | UD | 25.89 | 32.51 | UD | UD | 25.75 | 33.36 | UD | UD | UD | 25.25 | N | N | N |
| P55 | UD | UD | 23.41 | 31.87 | UD | UD | 23.54 | 32.70 | UD | UD | UD | 25.31 | N | N | N |
| P56 | UD | UD | 28.33 | 32.86 | UD | UD | 28.41 | 33.26 | UD | UD | UD | 25.66 | N | N | N |
| P57 | UD | UD | 28.34 | 32.90 | UD | UD | 28.40 | 33.24 | UD | UD | UD | 26.14 | N | N | N |
| P58 | 33.38 | 32.39 | 30.65 | 33.38 | 34.07 | 33.65 | 30.10 | 34.12 | 33.29 | 37.49 | 34.89 | 26.82 | P | P | P |
| P59 | 39.03 | UD | 28.94 | 32.99 | UD | UD | 28.64 | 33.57 | UD | UD | UD | 25.77 | N | N | N |
| P60 | 39.53 | UD | 27.94 | 31.77 | UD | UD | 27.67 | 32.46 | UD | UD | UD | 25.11 | N | N | N |
| P61 | UD | UD | 26.81 | 31.93 | UD | UD | 26.65 | 32.29 | UD | UD | UD | 24.90 | N | N | N |
| P62 | UD | UD | 32.13 | 36.32 | UD | UD | 31.71 | 36.92 | UD | UD | UD | 27.27 | N | N | N |
| P63 | UD | UD | 28.42 | 32.91 | UD | UD | 27.90 | 33.90 | UD | UD | UD | 25.95 | N | N | N |
| P64 | UD | UD | 31.67 | 34.94 | UD | UD | 31.54 | 35.91 | UD | UD | UD | 27.53 | N | N | N |
| P65 | UD | UD | 22.81 | 32.49 | UD | UD | 22.49 | 33.30 | UD | UD | UD | 25.98 | N | N | N |
| P66 | 29.86 | 29.45 | 22.54 | 31.03 | 30.72 | 30.51 | 22.53 | 31.77 | 29.45 | 30.68 | 30.76 | 24.86 | P | P | P |
| P67 | 32.66 | 31.37 | 24.95 | 31.89 | 32.65 | 32.39 | 25.25 | 32.77 | 32.25 | 34.90 | 33.81 | 25.78 | P | P | P |
| P68 | UD | UD | 24.51 | 32.57 | UD | UD | 24.28 | 33.37 | UD | UD | UD | 25.67 | N | N | N |
| P69 | UD | UD | 26.49 | 33.05 | UD | UD | 26.57 | 34.02 | UD | UD | UD | 25.48 | N | N | N |
| Patient | N1 | E | RPP30 | PhHV | N2 | E | RPP30 | PhHV | N | ORF1ab | S | MS2 | N1E-RP | N2E-RP | TaqPath |
| P70 | UD | UD | 24.67 | 31.73 | UD | UD | 24.43 | 32.22 | UD | UD | UD | 24.40 | N | N | N |
| P71 | 40.23 | UD | 20.35 | 30.96 | UD | UD | 20.32 | 31.66 | UD | UD | UD | 24.65 | N | N | N |
| P72 | 43.15 | UD | 22.83 | 32.07 | UD | UD | 22.59 | 32.43 | UD | UD | UD | 25.15 | N | N | N |
| P73 | UD | UD | 28.21 | 33.01 | UD | UD | 28.13 | 33.57 | UD | UD | UD | 25.95 | N | N | N |
| P74 | UD | UD | 25.78 | 32.16 | UD | UD | 25.66 | 32.53 | UD | UD | UD | 24.71 | N | N | N |
| P75 | 38.44 | 35.68 | 26.65 | 32.44 | 39.20 | 38.93 | 26.31 | 33.25 | 35.47 | UD | UD | 25.79 | P | P | Inc |
| P76 | 22.69 | 22.62 | 25.77 | 32.01 | 23.04 | 23.42 | 25.12 | 32.39 | 21.91 | 22.55 | 22.77 | 26.69 | P | P | P |
| P77 | 21.47 | 21.60 | 23.90 | 33.21 | 21.84 | 22.33 | 23.98 | 32.68 | 21.16 | 22.53 | 22.38 | 30.08 | P | P | P |
| P78 | UD | UD | 24.49 | 32.73 | UD | UD | 24.37 | 33.07 | UD | UD | UD | 25.72 | N | N | N |
| P79 | UD | UD | 27.14 | 33.85 | UD | UD | 27.06 | 35.00 | UD | UD | UD | 26.04 | N | N | N |
| P80 | UD | UD | 22.78 | 32.70 | UD | UD | 22.89 | 33.24 | UD | UD | UD | 25.80 | N | N | N |
| P81 | UD | UD | 23.01 | 32.61 | UD | UD | 22.74 | 32.79 | UD | UD | UD | 25.57 | N | N | N |
| P82 | UD | UD | 26.99 | 31.79 | UD | UD | 26.84 | 32.72 | UD | UD | UD | 25.52 | N | N | N |
| P83 | UD | UD | 31.29 | 35.55 | UD | UD | 31.42 | 35.57 | UD | UD | UD | 27.24 | N | N | N |
| P84 | UD | UD | 22.18 | 31.85 | UD | UD | 21.99 | 32.82 | UD | UD | UD | 24.75 | N | N | N |
| P85 | UD | UD | 24.37 | 32.50 | UD | UD | 24.14 | 33.25 | UD | UD | UD | 25.85 | N | N | N |
| P86 | UD | UD | 23.23 | 31.79 | UD | UD | 23.15 | 32.62 | UD | UD | UD | 24.80 | N | N | N |
| P87 | UD | UD | 23.10 | 31.79 | 44.55 | UD | 22.99 | 31.89 | UD | UD | UD | 24.76 | N | N | N |
| P88 | UD | UD | 24.51 | 32.62 | UD | UD | 24.48 | 33.49 | UD | UD | UD | 25.17 | N | N | N |
| P89 | UD | UD | 24.67 | 32.02 | UD | UD | 24.45 | 34.68 | UD | UD | UD | 25.17 | N | N | N |
| P90 | 25.58 | 25.96 | 23.32 | 31.65 | 26.04 | 26.56 | 23.27 | 32.18 | 24.15 | 24.71 | 24.87 | 24.73 | P | P | P |
| P91 | UD | UD | 20.27 | 31.55 | UD | UD | 20.30 | 31.207 | UD | UD | UD | 24.95 | N | N | N |
| P92 | 17.51 | 16.25 | 20.36 | 29.33 | 17.24 | 16.79 | 20.49 | 31.68 | 17.28 | 16.97 | 17.10 | 27.18 | P | P | P |
| P93 | UD | UD | 22.97 | 32.74 | UD | UD | 22.89 | 33.44 | UD | UD | UD | 25.60 | N | N | N |
| P94 | UD | UD | 23.60 | 32.46 | UD | UD | 23.70 | 33.29 | UD | UD | UD | 25.16 | N | N | N |
| P95 | UD | UD | 25.18 | 32.93 | UD | UD | 24.99 | 33.28 | UD | UD | UD | 26.64 | N | N | N |
| Patient | N1 | E | RPP30 | PhHV | N2 | E | RPP30 | PhHV | N | ORF1ab | S | MS2 | N1E-RP | N2E-RP | TaqPath |
| P96 | UD | UD | 23.53 | 33.50 | UD | UD | 23.31 | 34.08 | UD | UD | UD | 25.88 | N | N | N |
| P97 | UD | UD | 25.64 | 31.92 | UD | UD | 25.09 | 31.82 | UD | UD | UD | 24.83 | N | N | N |
| P98 | 21.50 | 20.39 | 23.62 | 31.84 | 21.29 | 20.90 | 23.20 | 32.92 | 20.73 | 20.43 | 21.33 | 25.97 | P | P | P |
| P99 | UD | UD | 23.01 | 32.16 | UD | UD | 23.43 | 34.11 | UD | UD | UD | 27.60 | N | N | N |
| P100 | 18.24 | 18.92 | 21.51 | 32.04 | 18.68 | 19.44 | 21.99 | 32.50 | 17.91 | 18.05 | 18.69 | 25.49 | P | P | P |
| P101 | UD | UD | 23.77 | 32.02 | UD | UD | 24.05 | 32.62 | UD | UD | UD | 25.25 | N | N | N |
| P102 | UD | UD | 24.71 | 30.80 | UD | UD | 24.59 | 31.29 | UD | UD | UD | 24.31 | N | N | N |
| P103 | UD | UD | 23.61 | 32.64 | UD | UD | 23.55 | 32.82 | UD | UD | UD | 25.13 | N | N | N |
| P104 | UD | UD | 23.12 | 31.92 | UD | UD | 22.77 | 32.44 | UD | UD | UD | 24.57 | N | N | N |
| P105 | 24.75 | 25.23 | 25.61 | 31.03 | 25.26 | 25.85 | 25.33 | 31.82 | 25.95 | 26.11 | 26.42 | 24.74 | P | P | P |
| P106 | UD | UD | 26.67 | 32.93 | UD | UD | 26.60 | 32.68 | UD | UD | UD | 25.36 | N | N | N |
| P107 | 33.90 | 32.43 | 21.76 | 31.97 | 34.09 | 33.04 | 21.71 | 31.89 | 32.29 | 33.97 | 32.74 | 24.80 | P | P | P |
| P108 | UD | UD | 27.70 | 32.01 | UD | UD | 27.35 | 33.17 | UD | UD | UD | 24.69 | N | N | N |
| **Controls** |  |  |  |  |  |  |  |  |  |  |  |  |  |  |  |
| -ve (extr) | UD | UD | UD | 31.75 | UD | UD | UD | 32.77 | UD | UD | UD | 25.18 | OK | OK | OK |
| -ve | UD | UD | UD | UD | UD | UD | UD | UD | UD | UD | UD | UD | OK | OK | OK |
| +ve | 30.89 | 34.15 | * | UD | 30.35 | 34.49 | * | UD | 31.28 | 31.88 | 31.23 | UD | OK | OK | OK |

UD, undetermined; P, positive; N, negative; Inc, inconclusive
-ve (extr), negative control with viral transport medium after RNA isolation (does not contain SARS-CoV-2 or human material; does contain PhHV);
-ve, negative control containing water only (should not contain any RNA)
+ve, positive control with in vitro transcribed RNA (50 copies; contains SARS-CoV-2 target RNA, does not contain human or PhHV RNA)
\* UD, but FAM-positive samples give ~5% signal bleed through into the JOE channel; this should be taken into account for samples without any true RPP30 (HEX) signal

**Table S5.** Percentage of known SARS-CoV-2 genomic sequences with mutations in primer/probe binding sites for RdRp, M and S gene assays.

| Assay | Primer/probe | Percentage of strains with any mismatch/deletion* |  | Percentage of strains with mismatch in 5 most 3' nt* |  |
| --- | --- | --- | --- | --- | --- |
|  |  | Per primer/probe | Per assay** | Per primer | Per assay*** |
| RdRp | RdRp_SARsR-F2 | 0.44% | 100% | 0.071% | 0.078% |
|  | RdRp_SARsR-P2 | 0.66% |  | (0.005%) |  |
|  | RdRp_SARsR-R1 | 100% |  | 0.007% |  |
| M gene | M-475-F | 0.16% | 1.11% | 0.042% | 0.045% |
|  | M-507-P | 0.87% |  | (0.857%) |  |
|  | M-574-R | 0.08% |  | 0.003% |  |
| S gene | S-Gene-F | 0.65% | 1.90% | 0.067% | 0.375% |
|  | S-Gene-P | 0.74% (0.31%)* |  | (0.043%) |  |
|  | S-Gene-R | 0.55% |  | 0.308% |  |

High quality genome sequences analysed only (n=97,782).
\* No genomes have deletions in primer/probe binding regions, with the exception of the S-gene probe, which has a 6 nt deletion in 0.31% of genome sequences. Presence of these changes does not necessarily impact on primer/probe performance. Mutations at the 3' ends for the primer regions (here defined as the final 5 nt) are more likely to affect assay sensitivity.
\*\* Mismatches/deletions in the primer/probe set used in single target assay
\*\*\* Mismatches in the forward and reverse primers only. For probes values are given between brackets for reference (in the "per primer" column).

**Table S6.** Interpretation and suggested action based on N1E-RP or N2E-RP qRT-PCR results.

| SARS-CoV-2 targets | RPP30 Cq | Interpretation | Suggested action |
| --- | --- | --- | --- |
| No positives<br>(UD, or Cq > 40) | High* | Inconclusive (possible negative), but poor sample | Repeat sampling |
|  | Low* | Negative | Report negative |
| One weak positive<br>(Cq 35-40) | High | Inconclusive; poor sample | Repeat sampling |
|  | Low | Inconclusive | Perform different assay with different primers/probes and of equal or higher sensitivity |
| Two weak positives<br>(Cq 35-40) | High | Positive; poor sample | Report positive |
|  | Low | Likely (weak) positive | Report positive |
| One moderate positive<br>(Cq 30-35) | High | Inconclusive, likely positive; poor sample | Perform different assay with different primers/probes and of equal or higher sensitivity (consider repeat sampling) |
|  | Low | Inconclusive, possible positive | Perform different assay with different primers/probes and of equal or higher sensitivity |
| Two moderate positives<br>(Cq 30-35) | High | Positive; poor sample, but does not affect conclusion | Report positive |
|  | Low | Positive | Report positive |
| One strong positive<br>(Cq < 30) | High | Inconclusive, likely positive; poor sample | Perform different assay with different primers/probes and of equal or higher sensitivity (consider repeat sampling) |
|  | Low | Inconclusive, likely positive (second target may have mutations) | Perform different assay with different primers/probes and of equal or higher sensitivity to rule out false positive |
| Two strong positives<br>(Cq < 30) | High | Positive; poor sample, but does not affect conclusion | Report positive |
|  | Low | Positive | Report positive |

High PhHV Cq values (>35; but depending on the exact amount used for spike-in; 2 to 3 cycles above the expected Cq is a good guide) indicate poor RNA isolation and/or presence of PCR inhibitors. Therefore, any negative (i.e. both SARS-CoV-2 targets not detected) or inconclusive samples (i.e. only one SARS-CoV-2 targets detected) should be interpreted in this context, and repeat sampling/testing must be performed. This has no impact on positive samples (both SARS-CoV-2 targets detected), which should still be reported as such.
\* The precise RPP30 Cq cut-off value remains to be determined. This requires collection of a much larger amount of data, including repeat sampling of sufficient numbers of patients with varying RPP30 Cq values and initial negative test results. As an initial guide repeat sampling could be performed for any sample with RPP30 Cq >30. For any samples with RPP30 Cq values undetermined or >40, repeat sampling is essential, unless both SARS-CoV-2 targets are <40, in which case the test result is positive.

## Supplementary Protocol 1: 4-plex SARS-CoV-2 RT-qPCR assay

### Primers/probes

Dissolve primers and probes (Table P1; HPLC purified) in 10 mM Tris, 0.1 mM EDTA, pH 8.0 (IDTE Cat. No. 11-05-01-09, or similar) for 100 μM stocks. Prepare primer/probe mixes (Tables P2-P7), and store aliquots at -20°C. Working stocks can be stored at 4°C.

**Table P1.**
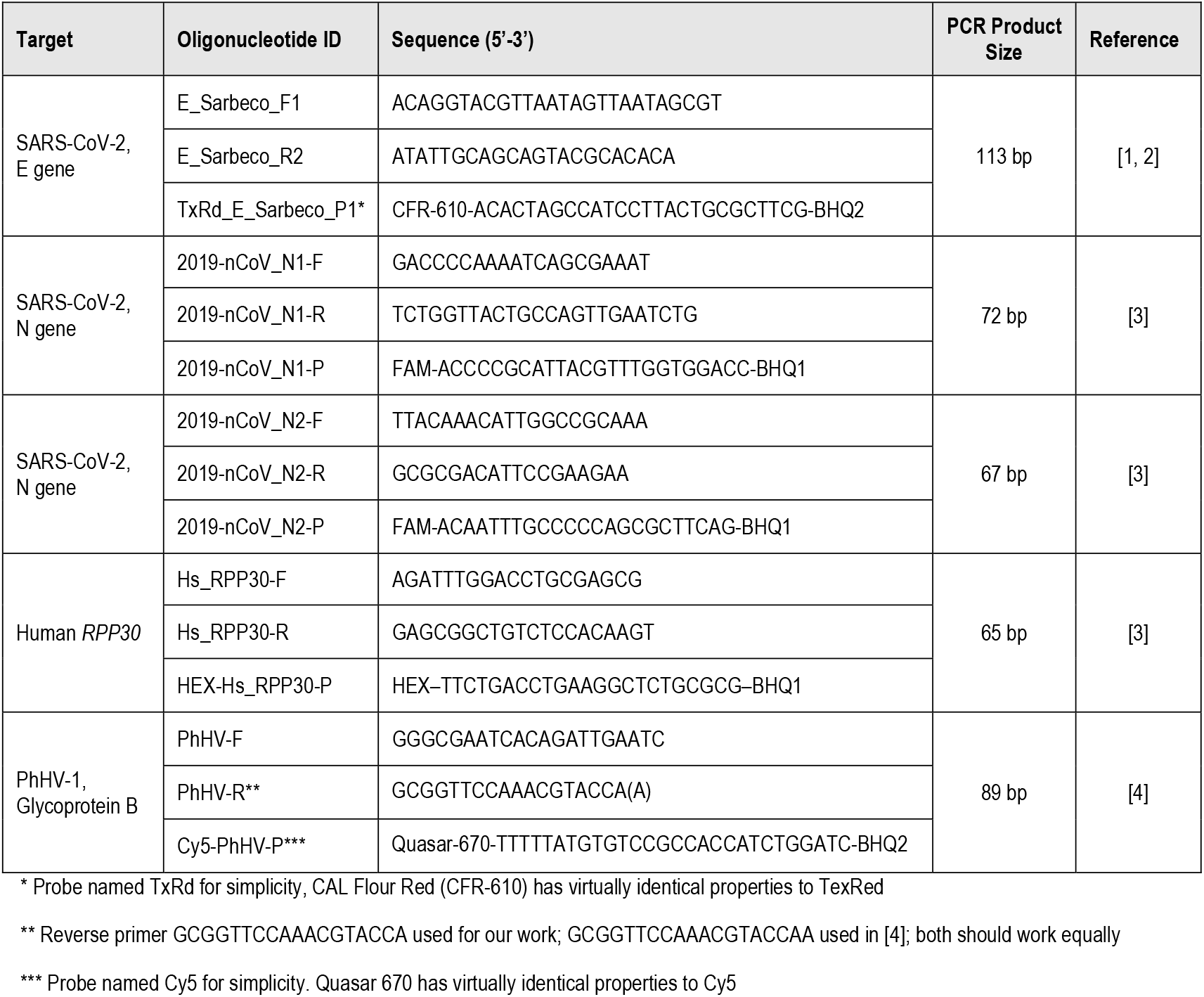
Primer/Probe details for 4-plex assays.

### Primer/Probe Mixes

**Table P2.**
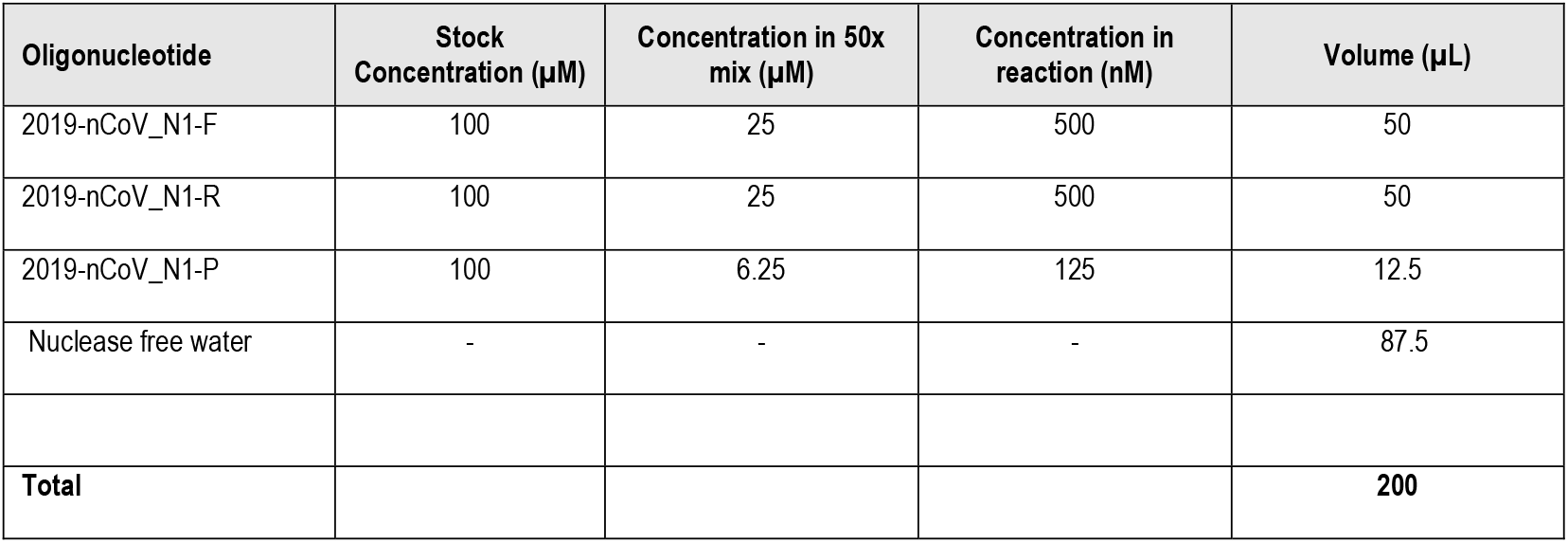
50x N1 (FAM) Primer/Probe Mix.

**Table P3.**
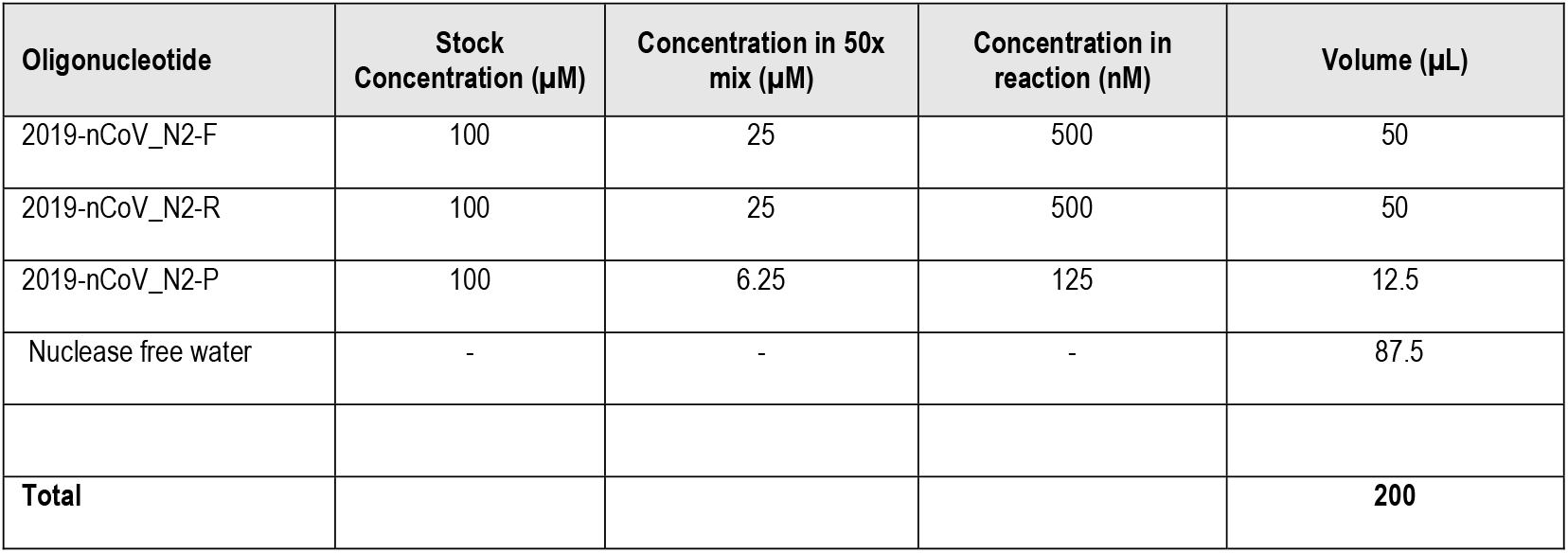
50x N2 (FAM) Primer/Probe Mix.

| Oligonucleotide | Stock Concentration (μM) | Concentration in 50x mix (μM) | Concentration in reaction (nM) | Volume (μL) |
| --- | --- | --- | --- | --- |
| 2019-nCoV_N2-F | 100 | 25 | 500 | 50 |
| 2019-nCoV_N2-R | 100 | 25 | 500 | 50 |
| 2019-nCoV_N2-P | 100 | 6.25 | 125 | 12.5 |
| Nuclease free water | - | - | - | 87.5 |
| **Total** |  |  |  | **200** |

**Table P4.** 50x E gene (TexRed) Primer/Probe Mix.

| Oligonucleotide | Stock Concentration (μM) | Concentration in 50x mix (μM) | Concentration in reaction (nM) | Volume (μL) |
| --- | --- | --- | --- | --- |
| E_Sarbeco_F1 | 100 | 20 | 400 | 40 |
| E_Sarbeco_R2 | 100 | 20 | 400 | 40 |
| TxRd_E_Sarbeco_P1 | 100 | 10 | 200 | 20 |
| Nuclease free water | - | - | - | 100 |
| **Total** |  |  |  | **200** |

**Table P5.** 50x *RPP30* (HEX) Primer/Probe Mix.

| Oligonucleotide | Stock Concentration (μM) | Concentration in 50x mix (μM) | Concentration in reaction (nM) | Volume (μL) |
| --- | --- | --- | --- | --- |
| Hs_RPP30-F | 100 | 25 | 500 | 50 |
| Hs_RPP30-R | 100 | 25 | 500 | 50 |
| HEX-Hs_RPP30-P | 100 | 6.25 | 125 | 12.5 |
| Nuclease free water | - | - | - | 87.5 |
| **Total** |  |  |  | **200** |

**Table P6.** 50x PhHV (Cy5) Primer/Probe Mix.

| Oligonucleotide | Stock Concentration (μM) | Concentration in 50x mix (μM) | Concentration in reaction (nM) | Volume (μL) |
| --- | --- | --- | --- | --- |
| PhHV-F | 100 | 15 | 300 | 30 |
| PhHV-R | 100 | 15 | 300 | 30 |
| Cy5-PhHV-P | 100 | 5 | 100 | 10 |
| Nuclease free water | - | - | - | 130 |
| **Total** |  |  |  | **200** |

**Table P7.** 12.5x 4-plex Primer/Probe Mix (for 100 reactions)

|  | Stock | Probe Concentration in 12.5x mix (μM) | Probe Concentration in reaction (nM) | Volume (μL) |
| --- | --- | --- | --- | --- |
| N1 or N2 mix (FAM) | 50x | 1.56 | 125 | 50 |
| E mix (TxRed) | 50x | 2.5 | 200 | 50 |
| *RPP30* mix (HEX) | 50x | 1.56 | 125 | 50 |
| PhHV mix (Cy5) | 50x | 1.25 | 100 | 50 |
| **Total** |  |  |  | **200** |

### RT-qPCR

Any Real-Time qPCR machine that can detect the four different channels and has been calibrated for the appropriate fluorophores can be used. We use the Applied Biosystems™ 7500 Fast Real-Time PCR Systems (channels FAM, JOE, TEXAS RED and CY5) and 7500 Software v2.3, MicroAmp Fast Optical 0.1mL 96-well reaction plates (Cat. No. 4346906) and Optical Adhesive film (Cat. No. 4311971). We performed all RT-qPCRs with Takara One Step PrimeScript™ III RT-qPCR kit (Cat. No. RR600B). Other One-Step mixes are available, but may require slightly different reaction conditions and may change sensitivity of SARS-CoV-2 detection.

### N1E-RP and N2E-RP 4-plex assay

Master Mix (20 μl per reaction), assemble on ice

|  |  |  |  |
| --- | --- | --- | --- |
| H<sub>2</sub>O (RNase free) | 5.5 µl | for 100x | 550 µl |
| 2x reaction mix | 12.5 µl |  | 1250 µl |
| 12.5x primer/probe mix | 2 µl |  | 200 µl |
| Mix with template RNA | 5 µl |  |  |

Spin down before transferring to PCR machine

### PCR program

|  |  |
| --- | --- |
| 52°C | 5 min (reverse transcription) |
| 95°C | 10 s (RT inactivation/denaturation) |
| 45 cycles of | 95°C 3 s (denaturation) |
|  | 60°C 30 s (amplification & detection) |

### Controls

Add the viral spike-in control to the lysis buffer master mix before sample inactivation.

[For our assays, we used 25 μl of culture supernatant containing PhHV particles to 25 ml of lysis buffer. This amount was previously shown to give a Cq value of ∼33.]

For each plate, include the following controls:

H10, negative extraction control, VTM extracted;

H11, non-extraction negative control, water only;

H12, 50 copies of positive control RNA (see below).

### Positive control RNA

Positive control RNAs generated by in vitro transcription (IVT) were provided by:

- Sylvie Behillil (Institut Pasteur, Paris, France): mix of E gene and RdRp (the latter is <u>not</u> the same as the Corman et al (2020) RdRp template) [1-3] at 10^9^ copies/μl.
- Christine Tait-Burkard (Roslin Institute, Edinburgh, UK): RdRp [1, 2], N1/N3 and N2 [3]. Three individual IVT RNAs at known concentrations (ng/μl) were provided; molecular weights used to determine the concentrations in copies/μl and 10^9^ copies/μl prepared for each RNA.

Prepare all stocks and dilutions in Eppendorf DNA LoBind tubes (Cat. No. 10051232).

1. Prepare an equimolar mix of all RNAs at 2.5 × 10^8^ copies/μl. Store aliquots of this solution at -80°C.
2. Prepare 10^4^ copies/μl positive RNA controls and store 5 μl aliquots at -80°C for single use per plate. Mix 5μl of 2.5 x 10^8^ copies/μl with 620 μl of water to give 2 × 10^6^ copies/μl. Dilute this 20 μl plus 180 μl water giving a 2 x 10^5^ copies/μl solution, and 20 μl plus 380 μl giving 10^4^ copies/μl.
3. For each plate, a 25 copies/μl solution is made by diluting the 10^4^ copies/μl solution by mixing:

> 2 μl with 98 μl water, then
>
> 12.5 μl of this with 87.5 μl water
>
> Of this 25 copies/μl solution, 2 μl is added to well H12 along with 3 μl of water, to give the 50 copy positive control on each plate.

## Supplementary Protocol 2: Viral nucleic acid isolation

In principle, the buffers and solution below can replace those of the equivalent buffers in the Omega Mag-Bind Viral DNA/RNA 96 Kit (Cat. No. M6246). Lysis and wash buffers can either be replaced with guanidine thiocyanate (GnSCN) or guanidine hydrochloride (GnHCl) containing solutions, depending on reagent availability. In our preliminary tests, all performed equally well (data not shown). All lysis buffers (Omega TNA, GnSCN Lysis Buffer, GnHCl Lysis Buffer, each with/without isopropanol) were shown to inactivate coronavirus after 15 min incubation (Fig P1). Briefly, to determine whether lysis buffers inactivate coronaviruses, 200 μl CoV 229E-GFP [5, 6] stock (9.6×10^5^ pfu/ml in DMEM, 10% FCS, 1% NEAA) was mixed with lysis buffer at the recommended ratio (240 μl lysis buffer without isopropanol or 520 μl lysis buffer with isopropanol, i.e. 240 μl buffer and 280 μl isopropanol). For positive infection controls, virus was mixed with 240 or 520 μl medium. All mixes were inverted 8 times and incubated at room temperature for 15 min. Cytotoxic components were then removed by centrifugation at 4°C using Microcon filter columns (Millipore; 30 kDa cut-off), and two 0.5 ml PBS washes, similar to previously described methods [7, 8]. Remaining virus particles were then resuspended in 200 μl DMEM and 50 μl of a 1/100 dilution added to HUH7 cells (a cell line permissive to infection by CoV 229E). Cells were seeded the previous day at 1.8×10^4^/well in a black 96-well plate (Corning), and were at ∼80% confluence for infection. Cultures were monitored daily for cell viability, cytopathic effects and GFP expression using microscopy. No significant cell death was observed for any of the samples. Relative fluorescence was measured using a Clariostar BMG Plate Reader at 72 h, with fluorescence for a non-infected control set to zero. No fluorescence was observed for any of the lysis buffer treated samples (Fig P1), and fluorescence microscopy confirmed the absence of GFP positive cells (data not shown), consistent with complete viral inactivation.

In our preliminary tests, we used the March 2020 version of the protocol provided with the Omega Mag-Bind Viral DNA/RNA 96 Kit to test viral nucleic acid isolation with our own solutions and reagents (see below). We used the Mag-Bind Particles CNR from the Omega kit, and although we have not yet tested this, we expect that these can be replaced by SeraSil-Mag silica beads (Cytiva, cat No. 29357375). The March 2020 protocol is different from the April 2020 Supplementary Protocol provided by Omega. The latter was used in combination with original Omega kit components for all other work presented in our manuscript. We do not see any reasons why our solutions would not work equally well with the April 2020 version of the protocol, but have not tested this. All purifications were carried out using a KingFisher Flex robot (Cat. No. 5400610), KingFisher Deep well plates (Cat. No. 95040450), KingFisher Flex 96 Deep-Well Tip Combs (Cat. No. A43074) and KingFisher 96 microplates (Cat. No. 97002540). Alternative robots could be used; and manual purifications are also possible.

**Figure P1.**
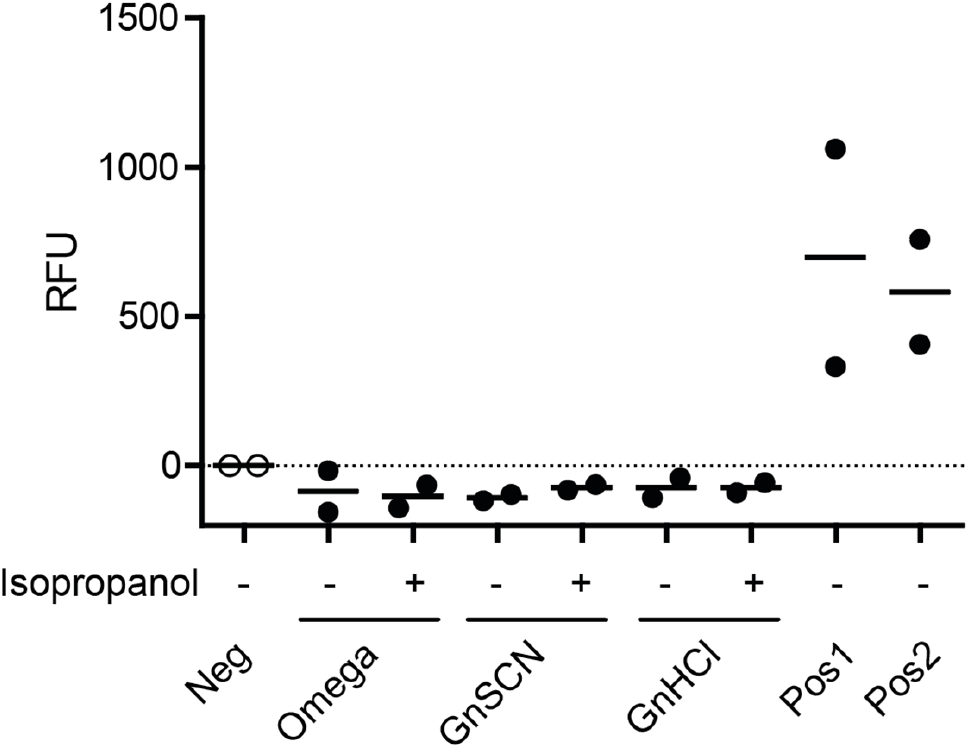
Inactivation of coronavirus CoV-229E by Omega TNA, and GnSCN and GnHCl lysis buffers. CoV 229E-GFP was mixed with lysis buffer (- or + isopropanol) as described and then used to infect HUH7 cells. After 72 h of infection, GFP fluorescence (indicating infected cells) was measured. Negative (Neg) control, no infection; positive control (Pos1 and 2), virus mixed with medium instead of lysis buffer. Fluorescence units (RFU) are expressed relative to background (negative control fluorescence set to 0). Solid lines indicate the mean for n = 2 independent experiments.

### Protocol in brief (tested with home-made solutions; based on Omega March 2020 protocol)

1. Per sample, prepare 528 μl master mix: 240 μl lysis buffer, 8 μl carrier RNA (1 μg/μl) and 280 μl isopropanol
2. Add 200 μl of patient sample in VTM, mix thoroughly; incubate for >15 min
3. Add 20 μl of 1:1 mix of Magnetic bead suspension and Proteinase K solution (40 μg/μl). KingFisher Flex, loop though 3 times: 2 min fast mix, 30 s half mix, 30 s bottom mix
4. Employ magnetic separation (Collect beads, 3×10 s)
5. Wash beads with 400 μL VHB wash buffer (Release beads, 3 min fast mix, collect beads 3×5 s)
6. Wash beads with 500 μL SPR Wash buffer (Release beads, 2 min fast mix, collect beads 3×5 s)
7. Wash beads with 500 μL SPR Wash buffer (Release beads, 2 min fast mix, collect beads 3×5 s)
8. Air dry magnetic beads (10 min, above well)
9. Elute nucleic acids in 50 μl nuclease-free water (Release beads, 5 min medium mix, collect beads 3×10 s)

### Protocol in brief (not tested with home-made solutions; based on Omega April 2020 protocol)

1. Per sample, mix 240 μl lysis buffer and 1 μl carrier RNA (1 μg/μl)
2. Add 200 μl of patient sample in VTM, mix thoroughly; incubate for >15 min
3. Add isopropanol bead mix (280 μl isopropanol and 2 μl magnetic beads). KingFisher Flex, loop though 3 times: 2 min fast mix, 30 s half mix, 30 s bottom mix
4. Employ magnetic separation (Collect beads, 3×10 s)
5. Wash beads with 350 μL VHB wash buffer (Release beads, 3 min fast mix, collect beads 3×5 s)
6. Wash beads with 350 μL SPR Wash buffer (Release beads, 2 min fast mix, collect beads 3×5 s)
7. Wash beads with 350 μL SPR Wash buffer (Release beads, 2 min fast mix, collect beads 3×5 s)
8. Air dry magnetic beads (10 min, above well)
9. Elute nucleic acids in 50 μl nuclease-free water (Release beads, 5 min medium mix, collect beads 3×10 s)

## Reagents

### Guanidine thiocyanate based

#### GnSCN Lysis Buffer (TNA Lysis buffer equivalent)

|  | Amount for 1 L | Concentration |
| --- | --- | --- |
| Guanidine thiocyanate | 473 g | 4 M |
| Sarkosyl (sodium lauroyl sarcosinate) | 20 g | 2% |
| 1 M Tris-HCl pH 7.5 | 50 ml | 50 mM |
| 0.5 M EDTA pH 8.0 | 20 ml | 10 mM |
| β-mercaptoethanol | 10 ml | 1% |

#### GnSCN Wash buffer 1 (VHB Wash buffer equivalent)

|  | Amount for 1 L | Concentration |
| --- | --- | --- |
| Guanidine thiocyanate | 118 g | 1 M |
| 1 M Sodium citrate pH 7* | 10 ml | 10 mM |
| Ethanol | 560 ml | 56% |

\*to set pH, add 10 ml 1 M HCl for every 500 ml 1 M Sodium citrate.

### Guanidine hydrochloride based

#### GnHCl Lysis Buffer (TNA Lysis buffer equivalent)

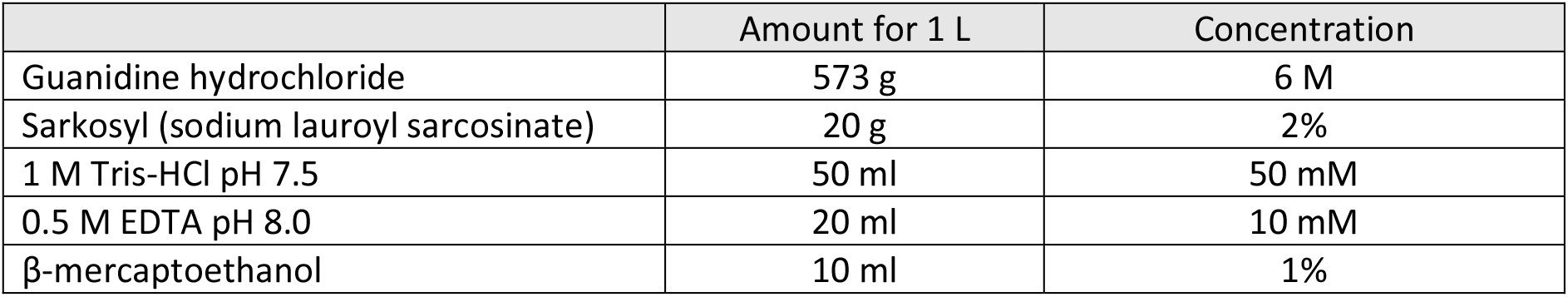

#### GnHCl Wash Buffer 1 (VHB Wash buffer equivalent)

|  | Amount for 1 L | Concentration |
| --- | --- | --- |
| Guanidine hydrochloride | 107 g | 1.25 M |
| 1 M Sodium citrate pH 7* | 10 ml | 10 mM |
| Ethanol | 560 ml | 56% |

\*to set pH, add 10 ml 1 M HCl for every 500 ml 1 M Sodium citrate.

### SPR Wash buffer 2 equivalent

80% Ethanol

### Proteinase K (PCR grade; e.g. Roche Cat. No. 03115801001)

Dissolve in 10 mM Tris pH 8.0, 1 mM EDTA at 40 mg/ml Store aliquots at -20°C

### Carrier RNA (e.g. yeast tRNA, Roche Cat. No. 10109509001)

Dissolve in 10 mM Tris pH 8.0, 1 mM EDTA at 1 mg/ml Store aliquots at -20°C

